# Outpatient regimens to reduce COVID-19 hospitalisations: a systematic review and meta-analysis of randomized controlled trials

**DOI:** 10.1101/2022.05.24.22275478

**Authors:** David J. Sullivan, Daniele Focosi, Daniel F. Hanley, Mario Cruciani, Massimo Franchini, Jiangda Ou, Arturo Casadevall, Nigel Paneth

## Abstract

**Background:** During pandemics, early outpatient treatments reduce the health system burden. Randomized controlled trials (RCTs) in COVID-19 outpatients have tested therapeutic agents, but no RCT or systematic review has been conducted comparing the efficacy of the main outpatient treatment classes to each other. We aimed in this systematic review of outpatient RCTs in COVID-19 to compare hospitalisation rate reductions with four classes of treatment: convalescent plasma, monoclonal antibodies, small molecule antivirals and repurposed drugs.

**Methods:** We conducted a systematic review and meta-analysis of all COVID-19 outpatient RCTs that included the endpoint of progression to hospitalisation. We assembled, from multiple published and preprint databases, participant characteristics, hospitalisations, resolution of symptoms and mortality from January 2020 to May 21, 2023. The risk of bias from COVID-NMA was incorporated into the Grading of Recommendations Assessment, Development and Evaluation (GRADE) system. We measured heterogeneity with I^2^. Meta-analysis by a random or fixed effect model dependent on significant heterogeneity (I^2^ >50%) was performed. The protocol was registered in PROSPERO, CRD42022369181.

**Findings:** The search identified 281 studies of which 54 RCTs for 30 diverse interventions were included in the final analysis. These trials, performed largely in unvaccinated cohorts during pre-Omicron waves, focused on populations with at least one COVID-19 hospitalisation risk factor. Grouping by class, monoclonal antibodies (OR=0.31 [95% CI=0.24-0.40]) had highest efficacy, followed by COVID-19 convalescent plasma (CCP) (OR=0.69 [95% CI=0.53 to 0.90]) and small molecule antivirals (OR=0.78 [95% CI=0.48-1.33]) for hospital reduction. Repurposed drugs (OR=0.82 [95% CI-0.72-0.93]) had lower efficacy.

**Interpretation:** Inasmuch as omicron sublineages (XBB and BQ.1.1) are now resistant to monoclonal antibodies, oral antivirals are the preferred treatment in outpatients where available, but intravenous interventions from convalescent plasma to remdesivir are also effective and necessary in constrained medical resource settings or for acute and chronic COVID-19 in the immunocompromised.

**Funding:** US Department of Defense and National Institute of Health

**Research in context:** *Evidence before this study:* We systematically searched the published and preprint data bases for outpatient randomized clinical trials of treatment of COVID-19 disease with hospitalisation as an endpoint. Previous systematic reviews and meta-analyses have confined the reviews to specific classes such as convalescent plasma, monoclonal antibodies, small molecule antivirals or repurposed drugs. Few comparisons have been made between these therapeutic classes. The trials took place both in the pre-vaccination and the vaccination era, spanning periods with dominance of different COVID variants. We sought to compare efficacy between the four classes of treatments listed above when used in outpatient COVID-19 patients as shown in randomized, placebo-controlled trials.

**Added value of this study:** This systematic review and meta-analysis brings together trials that assessed hospitalisation rates in diverse COVID-19 outpatient populations varying in age and comorbidities, permitting us to assess the efficacy of interventions both within and across therapeutic classes. While heterogeneity exists within and between these intervention classes, the meta-analysis can be placed in context of trial diverse populations over variant time periods of the pandemic. At present most of the world population has either had COVID-19 or been vaccinated with a high seropositivity rate, indicating that future placebo-controlled trials will be limited because of the sample sizes required to document hospitalisation outcomes.

**Implications of all the available evidence:** Numerous diverse therapeutic tools need to be ready for a resilient response to changing SARS-CoV-2 variants in both immunocompetent and immunocompromised COVID-19 outpatient populations. To date few head-to-head randomized controlled trials (RCTs) has compared treatment options for COVID-19 outpatients, making comparisons and treatment choices difficult. This systematic review compares outcomes among RCTs of outpatient therapy for COVID-19, taking into account time between onset of symptoms and treatment administration. We found that small-chemical antivirals, convalescent plasma and monoclonal antibodies had comparable efficacy between classes and amongst interventions within the four classes. Monoclonals have lost efficacy with viral mutation, and chemical antivirals have contraindications and adverse events, while intravenous interventions like convalescent plasma or remdesivir remain resilient options for the immunocompromised, and, in the case of CCP, in resource constrained settings with limited availability of oral drugs.

## INTRODUCTION

By May 17, 2023 the world had recorded over 766 million cases and more than 6.9 million deaths from COVID-19. In the US, some 100 million cases have been recorded, with over a million deaths, while six million hospital admissions for COVID took place between August 2020 and December 2022. A pronounced spike in hospitalisations for COVID-19 in the US took place in the first two months of 2022 with the arrival of the Omicron variant of concern (VOC). Several approaches to reducing the risk of hospitalisation have been taken during the pandemic, including administering COVID-19 convalescent plasma (CCP), monoclonal antibodies (mAbs), small molecule antivirals or repurposed drugs. Vaccination and boosters have substantially reduced the hospitalisation and death risk, but outpatients at elevated severe COVID risk can still benefit from early treatment to avoid hospitalisation. Randomized controlled trials (RCTs) in outpatients have tested therapeutic agents against placebo or standard of care, but very few RCTs has been conducted that compare the main outpatient treatment classes.

The first outpatient treatments for COVID-19 authorized by the FDA were for mAbs (bamlanivimab, bamlanivimab plus etesevimab^1^ or casirivimab plus imdevimab^2^), approvals that preceded the introduction of mRNA vaccines^3, 4^. While many small molecules were repurposed as antivirals during the early stages of the pandemic, oral antivirals developed against SARS-CoV-2 for outpatients were not authorized and available until December 2021, when nirmatrelvir/ritonavir^5^ and molnupiravir^6^ were approved. The following month, intravenous remdesivir was also approved for outpatient use^7^. On December 2021, nearly two years after the first use of CCP, the FDA approved CCP outpatient use, but only for immunocompromised patients^8, 9^. The mAbs have been withdrawn due to viral variants BQ.1.* and XBB.* resistance. Here we assemble outpatient RCTs of COVID-19 different therapies, sharing either all cause or COVID-19 related hospitalisation for an endpoint. A literature search of MEDLINE (through PubMed), medRxiv and bioRxiv databases was carried out aiming to include all such RCTs published from March 2020 to October 2022 and these are summarized in the PRISMA chart (Figure 1). This systematic review and meta-analysis of outpatient COVID-19 RCTs, compared outcomes amongst CCP, mAbs, antivirals or repurposed drugs taking into account risk factors for progression, intervention dosage, time between symptom onset and treatment administration, and predominant variants of concern during RCTs.

**Figure 1.**
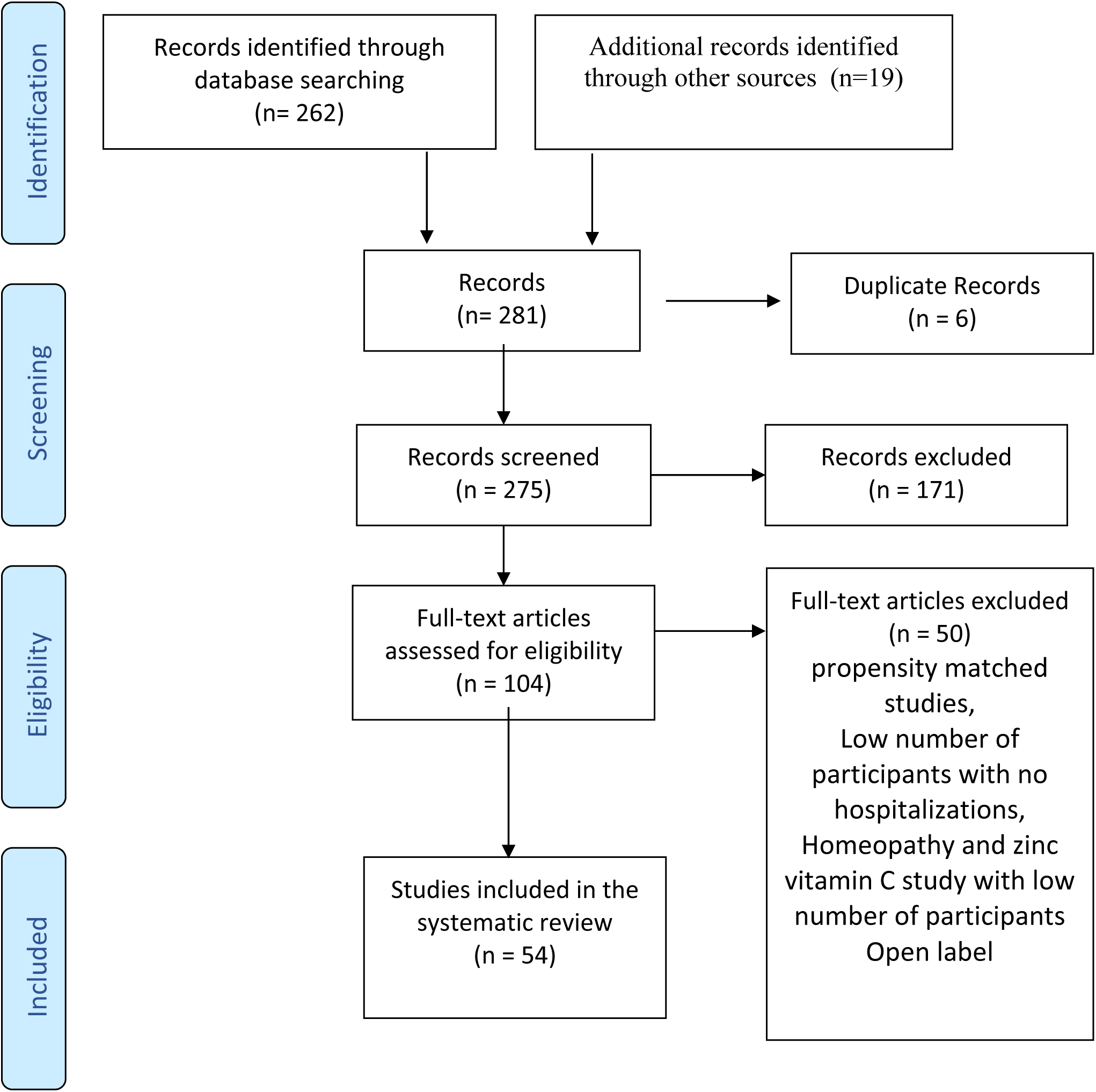
PRISMA flowchart for randomized controlled trials (RCT) selection in this systematic review.

## METHODS

The protocol has been registered in PROSPERO, the prospective register of systematic reviews and meta-analyses of the University of York (protocol registration number CRD42022369181). We assembled outpatient COVID-19 RCTs with hospitalisation or a single CCP study with life-threatening respiratory distress^10^ as the outcome, by searching MEDLINE (through PubMed), medRxiv and bioRxiv databases for the period of March 1, 2020 to May 22, 2022, with English language as the only restriction. The Medical Subject Heading (MeSH) and search query used were: “(“COVID-19” OR “SARS-CoV-2” OR “coronavirus disease 2019”) AND (“treatment” OR “therapy”) AND (“outpatient”) AND (“hospitalisation”)” AND (“randomized clinical trial”). We also screened the reference list of reviewed articles for studies not captured in our initial search. We excluded case reports, case series, retrospective, propensity-matched studies, non-randomized clinical trials, review articles, meta-analyses, studies with fewer than 30 participants, studies that did not record or had no hospitalisations. and articles reporting only aggregate data. Trials of COVID prevention^11^ were excluded, even if hospitalisations were recorded. Articles underwent a blind evaluation for inclusion by two assessors (D.S. and D.F.) and disagreements were resolved by a third senior assessor (A.C.). Figure 1 shows a PRISMA flowchart of the literature reviewing process. The following parameters were extracted from studies: baseline SARS-CoV-2 serology status, time from onset of symptoms to treatment, study dates, recruiting countries, gender, age (including the fraction of participants over age 50, 60 and 65), ethnicity, risk factors for COVID-19 progression (systemic arterial hypertension, diabetes mellitus, and obesity), sample size, dosage type of control, hospitalisations and deaths in each arm, and time to symptom resolution. Study dates were used to infer predominant VOCs.

### Assessment of risk of bias and GRADE assessment

A risk of bias assessment of each selected RCT was performed by COVID-19-NMA initiative^12, 13^. Within-trial risk of bias was assessed, using the Cochrane ROB tool for RCTs^14^.We explored clinical heterogeneity (e.g., risk factors for progression, time between onset of symptoms and treatment administration, and predominant variants of concern at the time of the interventions) and calculated statistical heterogeneity using τ², Cochran’s Q and estimated this using the I² statistic, which examines the total variation percentage across studies due to heterogeneity rather than chance.

We used the GRADE (The Grading of Recommendations Assessment, Development and Evaluation) system criteria to assess the quality of the body of evidence associated with specific outcomes, and constructed a ‘Summary of Findings’ table (Appendix Table 3), which defines the certainty of a body of evidence as the extent to which one can be confident that an estimate of effect or association is close to the true quantity of specific interest^14^. Publication bias was assessed by visual inspection of funnel plots.

### Statistical methods

Descriptive analysis included time-to-treatment, geography (country) of the study, age, sex, race, ethnicity, seropositive, hospital type and medical high-risk conditions. The unweighted pooling ARR, RRR, NNT were calculated based on the arithmetic summation of the total hospitalisation or death numbers in each therapeutic category.

Odds ratios (OR) and 95% confidence intervals (CI) were used to show the direction of effect and its significance in comparing treatment group and control groups. The studies were weighted with the Mantel-Haenszel method. The effect heterogeneity was calculated estimating the I-squared (I^2^) inconsistency index. If significant heterogeneity was detected (I^2^ > 50% and Cochran’s Q test for heterogeneity was significant (p < 0.10)), a random effect model was performed; otherwise, a fixed (common) effect model was performed. Weight, heterogeneity, between-study variance, and significance level were displayed in forest plots.

The significance level was 0.05. The figures were created in Prism software, R (version 4.2.1) and its statistical package “meta” (version 6.0-0). All the data manipulation and the analyses were performed in Excel, Prism, MedCalc (version 20.106), R and REVMAN.5.

Role of the funding source The study sponsors did not contribute to the study design; to the collection, analysis, and interpretation of data; to manuscript preparation, nor to the decision to submit the paper for publication.

## RESULTS TRIAL CHARACTERISTICS

We reviewed in detail 54 distinct outpatient RCTs (30 different interventions), conducted from March 2020 to May 22, 2023, across waves sustained by different SARS-CoV-2 variants of concern (VOC) and different vaccination periods in diverse patient populations (Figure 2). The studies varied in reporting outcomes of hospitalisation, whether all-cause or COVID-19 related. The CCP group included 3 RCTs with all-cause hospitalisation, 2 trials with COVID-19 related hospitalisations only and one trial with life-threatening respiratory distress in elderly individuals, deemed equivalent to hospital outcome (Appendix Table 1). The mAb RCTs included 3 trials with all-cause hospitalisations and 5 that used COVID-19 related hospitalisations as the outcome. The small molecule antivirals had 6 RCTs with all-cause hospitalisations and 5 with COVID-19 related hospitalisations, while 17 RCT’s of repurposed drugs used all-cause hospitalisations and 10 trials restricted to COVID-19 related hospitalisations. Here we report the hospitalisations as a composite of the hospital types. Because inclusion criteria varied across the RCTs, the power to detect a difference in hospitalisation rates varied across studies. Three CCP RCTs had higher control arm hospitalisation rates (11% – 31%) than all other antiviral RCTs, indicating that they studied sicker populations.^9^ (Table 1 and Figure 3). The six mAb RCTs had hospitalisation rates in the controls of 4.6-8.9%, the same range as CSSC-004^9^ (6.3%). Control hospitalisation rates in the molnupiravir-MOVE-OUT^7^, nirmatrelvir/ritonavir^5^ and remdesivir^15^ RCTs ranged from 5.3% to 9.7%. Low hospitalisation rates were found in RCTs that had many vaccinees (metformin-COVID-OUT – 3.2%^16^) or in which most participants were seropositive (molnupiravir-PANORAMIC – 0.8%). Low control arm hospitalisation rates were also found in two mAb RCTs – the bebtelovimab trial (1.6%)^17^ and REGN-CoV phase 1/2 (<2%), with the bebtelovimab RCT focusing on low-risk patients ^17^

**Figure 2.**
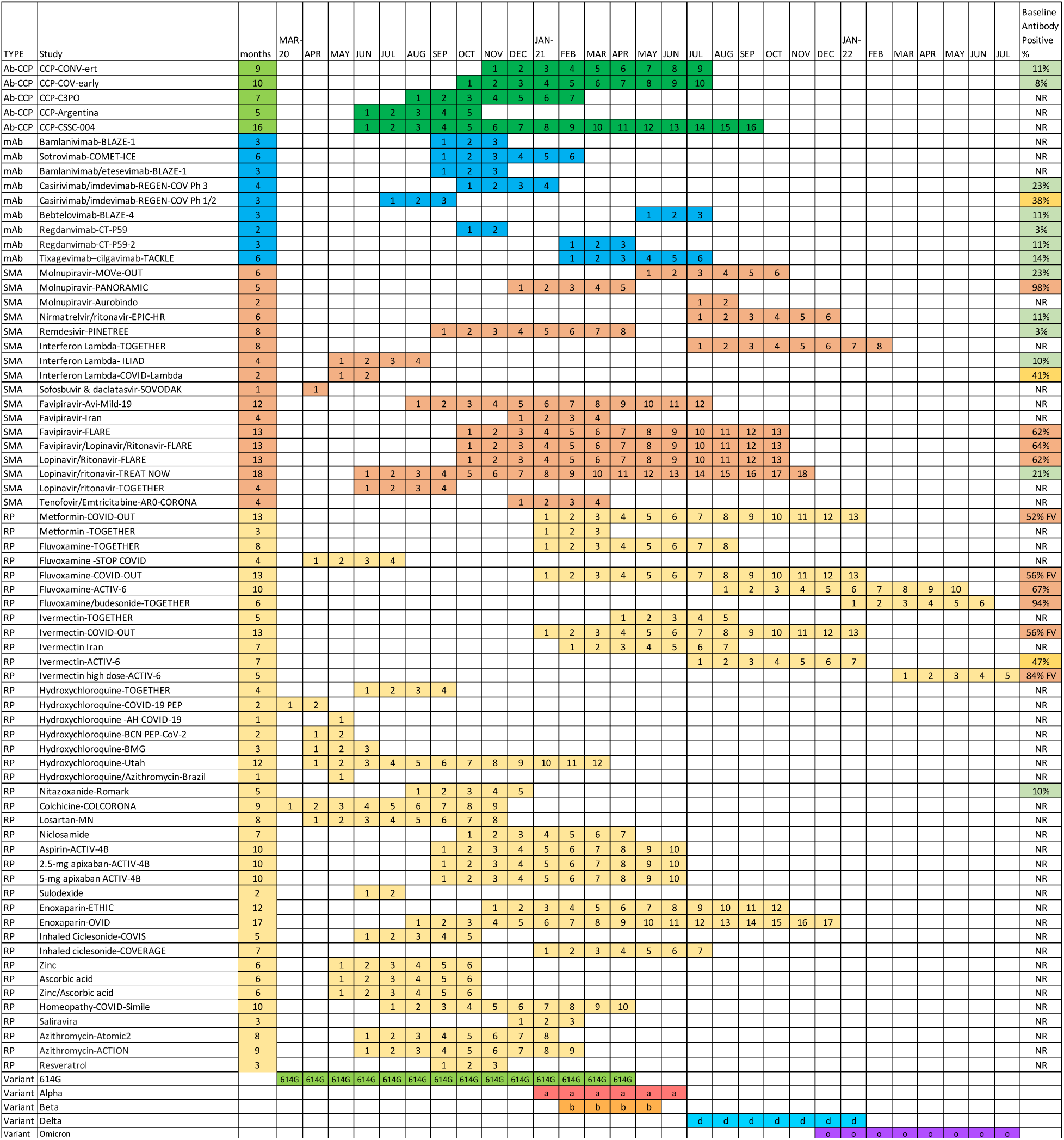
Duration and calendar months of the RCT in context of dominant variant(s) of concern and seropositivity rates. Study start and end for enrollments are charted with approximate time periods for variants of concern.

**Figure 3.**
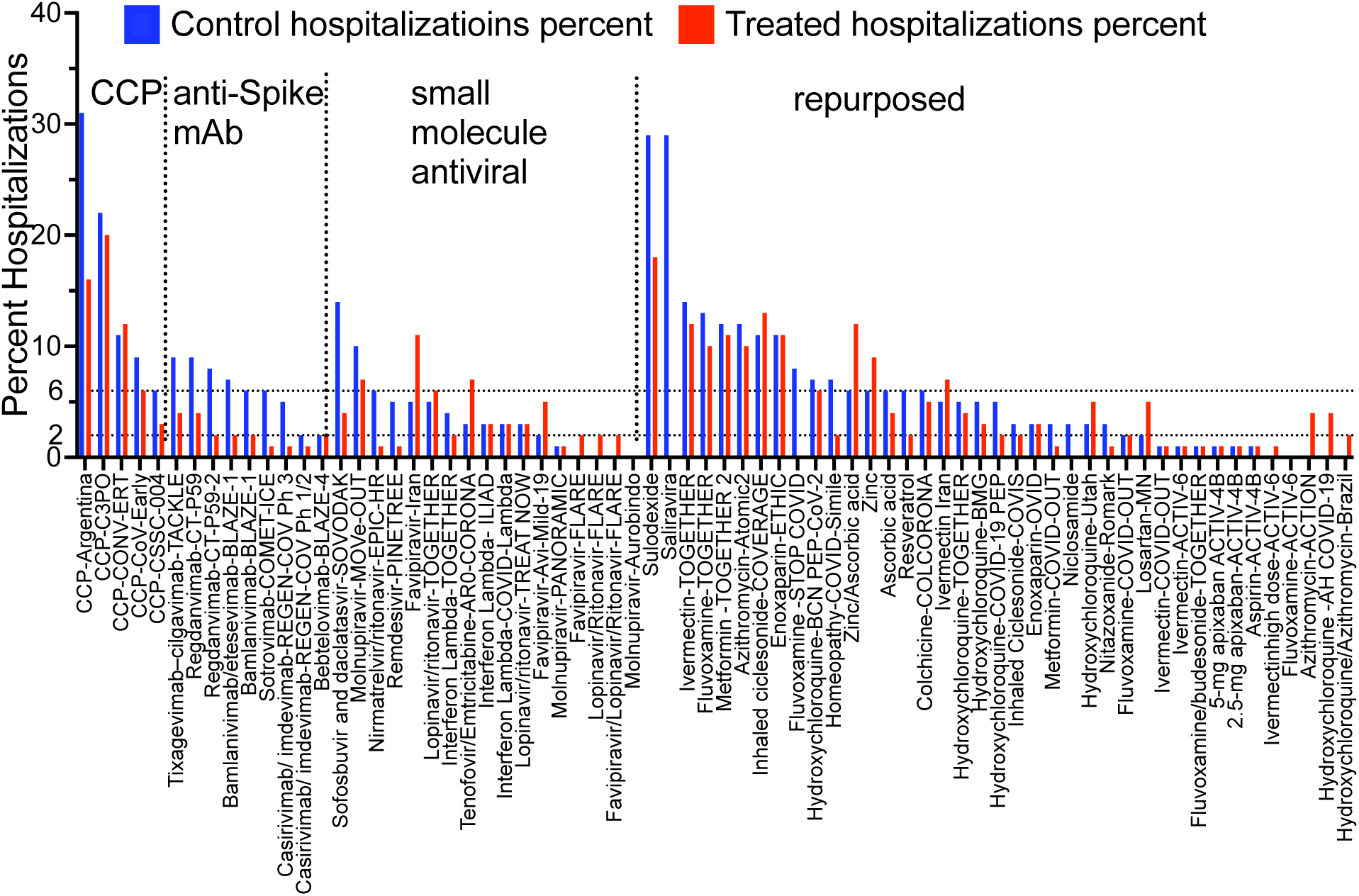
Percent hospitalisations in control groups sorted by therapy type and descending control hospitalisation rates.

**Table 1.** Hospital rates, risk reductions, NNT, numbers and symptom resolution.

| Study | Control hospitalizations % | hospitalizations % in intervention arm | ARR percent (95% CI) | RRR percent (95% CI) | NNT to prevent 1 hospitalization | Hospitalization (n) in control arm | total pts in control arm (n) | Hospitalization (n) in intervention arm | Total pts (n) in intervention arm | Symptom resolution: median duration- Intervention to control in days |
| --- | --- | --- | --- | --- | --- | --- | --- | --- | --- | --- |
| CCP (5 RCT) % or totals | 12 | 8.8 | 3.2 (0.9, 5.6) | 26.8 (8.1, 41.7) | 31 | 158 | 1315 | 116 | 1319 |  |
| anti-Spike mAbs (8 RCT) % or totals | 5.9 | 1.9 | 4.0 (3.2, 4.8) | 67.8 (59.1, 74.7) | 25 | 242 | 4102 | 88 | 4634 |  |
| Small molecule antiviral (11 RCTs) total or average | 1.8 | 1.3 | 0.5 (0.3, 0.8) | 29.1 (15.9, 40.2) | 187 | 314 | 17079 | 222 | 17025 |  |
| Small molecule antiviral (10 RCTs - w/o Mol-Pan.) total or average | 4.7 | 2.6 | 2.1 (1.3, 2.9) | 44.4 (30.7, 55.3) | 48 | 218 | 4595 | 119 | 4509 |  |
| Repurposed drugs (20 RCTs) total or average | 5.3 | 4.2 | 1.1 (0.5, 1.6) | 20.1 (10.1, 28.9) | 94 | 590 | 11121 | 483 | 11391 |  |
| All (47 RCTs) total or average | 3.9 | 2.6 | 1.2 (1.0, 1.5) | 31.8 (25.9, 37.3) | 81 | 1304 | 33617 | 909 | 34369 |  |
| CCP-CONV-ert<sup>24</sup> | 11.2 | 11.7 | -0.5 (-7.0, 5.9) | -4.8 (-83.9, 40.3) | -188 | 21 | 188 | 22 | 188 | NO difference 12 d vs 12 d |
| CCP-COV-Early<sup>35</sup> | 9.3 | 5.9 | 3.4 (-1.8, 8.5) | 36.2 (-27.9, 68.2) | 29 | 19 | 204 | 12 | 202 | NO difference 13 d vs 12 d |
| CCP-C3PO<sup>36</sup> | 22.0 | 20.2 | 1.8 (-5.3, 8.9) | 8.2 (-28.3, 34.4) | 55 | 56 | 254 | 52 | 257 | NO difference |
| CCP-Argentina<sup>10</sup> | 31.3 | 16.3 | 15.0 (2.0, 28.0) | 48.0 (5.8, 71.3) | 7 | 25 | 80 | 13 | 80 | Not reported |
| CCP-CSSC-004<sup>9</sup> | 6.3 | 2.9 | 3.4 (1.0, 5.8) | 54.3 (19.7, 74.0) | 29 | 37 | 589 | 17 | 592 | Not reported |
| CCP-Argentina (high titer)<sup>10</sup> | 31.3 | 8.3 | 22.9 (9.3, 36.5) | 73.3 (17.4, 91.4) | 4 | 25 | 80 | 3 | 36 | Not reported |
| CCP-CSSC-004 (<= 5 days)<sup>9</sup> | 9.7 | 1.9 | 7.7 (3.7, 11.7) | 79.9 (48.4, 92.2) | 13 | 25 | 259 | 5 | 257 | Not reported |
| Bamlanivimab-BLAZE-1<sup>1</sup> | 6.3 | 1.6 | 4.7 (0.5, 8.9) | 74.3 (24.7, 91.2) | 21 | 9 | 143 | 5 | 309 | NO difference 11 d to 11 d |
| Sotrovimab-COMET-ICE<sup>37</sup> | 5.7 | 1.1 | 4.5 (2.4, 6.7) | 80.0 (52.3, 91.6) | 22 | 30 | 529 | 6 | 528 | Not reported |
| Bamlanivimab/etesevimab-BLAZE-1<sup>25</sup> | 7.0 | 2.1 | 4.8 (2.3, 7.4) | 69.5 (40.8, 84.3) | 21 | 36 | 517 | 11 | 518 | YES- 8d vs 9d p=0.007 |
| Casirivimab/imdevimab-REGEN-COV Ph 3<sup>2</sup> | 4.6 | 1.3 | 3.3 (2.0, 4.6) | 71.3 (51.7, 82.9) | 30 | 62 | 1341 | 18 | 1355 | YES- 10 d vs 14 p=0.0001 |
| Casirivimab/imdevimab-REGEN-COV Ph 1/2<sup>38</sup> | 1.9 | 0.6 | 1.3 (-0.4, 3.1) | 70.1 (-24.4, 92.8) | 76 | 5 | 266 | 3 | 533 | Not reported |
| Bebtelovimab-BLAZE-4<sup>17</sup> | 1.6 | 1.6 | -0.04 (-3.1, 3.0) | -2.4 (-615.7, 85.4) | -2667 | 2 | 128 | 2 | 125 | YES- 6d to 8d p=0.003 |
| Regdanvimab-CT-P59<sup>26</sup> | 8.7 | 4.4 | 4.2 (-1.9, 10.3) | 48.8 (-25.2, 79.0) | 23 | 9 | 104 | 9 | 203 | YES 6 d vs 9 d p=0.01 |
| Regdanvimab-CT-P59-2<sup>27</sup> | 7.9 | 2.4 | 5.4 (3.1, 7.8) | 69.1 (46.4, 82.2) | 18 | 52 | 659 | 16 | 656 | 8 d to 13 d |
| Tixagevimab-cilgavimab-TACKLE<sup>39</sup> | 8.9 | 4.4 | 4.5 (1.1, 7.9) | 50.4 (14.3, 71.3) | 22 | 37 | 415 | 18 | 407 | Not reported |
| Molnupiravir-MOVE-OUT<sup>6</sup> | 9.7 | 6.8 | 3.0 (0.1, 5.8) | 30.4 (0.8, 51.2) | 34 | 68 | 699 | 48 | 709 | NO difference |
| Molnupiravir-PANORAMIC<sup>40</sup> | 0.8 | 0.8 | -0.1 (-0.3, 0.2) | -7.0 (-41.2, 18.9) | -1853 | 96 | 12484 | 103 | 12516 | YES 9 d vs 15 d |
| Molnupiravir-Aurobindo<sup>41</sup> | 0.0 | 0.0 | NC | NC | 0 | 0 | 610 | 0 | 610 | Yes 10 d vs 14 d p<0.001 |
| Nirmatrelvir/ritonavir-EPIC-HR<sup>5</sup> | 6.3 | 0.8 | 5.5 (4.0, 7.1) | 87.8 (74.7, 94.1) | 18 | 66 | 1046 | 8 | 1039 | Not reported |
| Remdesivir-<br>PINETREE<sup>15</sup> | 5.3 | 0.7 | 4.6 (1.8,<br>7.4) | 86.5 (41.4,<br>96.9) | 22 | 15 | 283 | 2 | 279 | YES- Alleviation of<br>symptoms by day 14<br>(rate ratio, 1.92; 95%<br>CI, 1.26 to 2.94) |
| Interferon Lambda-TOGETHER<sup>42</sup> | 3.9 | 2.3 | 1.7 (0.1,<br>3.2) | 42.6 (3.4,<br>65.9) | 60 | 40 | 1018 | 21 | 931 |  |
| Interferon Lambda-ILIAD<sup>43</sup> | 3.3 | 3.3 | 0 (-9.1,<br>9.1) | 0 (-1426,<br>93.4) |  | 1 | 30 | 1 | 30 | No difference |
| Interferon Lambda-COVID-Lambda<sup>44</sup> | 3.3 | 3.3 | 0 (-6.4,<br>6.4) | 0 (-586.9,<br>85.4) |  | 2 | 60 | 2 | 60 | NO difference 20 d<br>vs 20 d |
| Sofosbuvir and<br>daclatasvir-SOVODAK<sup>45</sup> | 14.3 | 3.7 | 10.6 (-<br>4.2, 25.4) | 74.1 (-117,<br>96.9) | 9 | 4 | 28 | 1 | 27 | NO difference in 7 d<br>symptoms |
| Favipavir-Avi-Mild-19<sup>46</sup> | 1.7 | 5.4 | -3.7 (-<br>8.4, 1.1) | -219 (-1447,<br>34.3) | -27 | 2 | 119 | 6 | 112 | NO difference 7d vs<br>7d |
| Favipiravir-Iran<sup>47</sup> | 5.1 | 10.5 | -5.4 (-<br>17.4, 6.6) | -105.3 (-<br>955.6, 60.1) | -19 | 2 | 39 | 4 | 38 |  |
| Favipiravir-FLARE<sup>48</sup> | 0.0 | 1.7 | -1.7 (-<br>5.0, 1.6) | NA | -59 | 0 | 60 | 1 | 59 |  |
| Favipiravir/Lopinav<br>ir/Ritonavir-FLARE<sup>48</sup> | 0.0 | 1.6 | -1.6 (-<br>4.8, 1.5) | NA | -61 | 0 | 60 | 1 | 61 |  |
| Lopinavir/Ritonavir<br>-FLARE<sup>48</sup> | 0.0 | 1.7 | -1.7 (-<br>5.0, 1.6) | NA | -60 | 0 | 60 | 1 | 60 |  |
| Lopinavir/ritonavir-TREAT NOW<sup>49</sup> | 2.7 | 3.2 | -0.5 (-<br>3.7, 2.6) | -19.8 (-251,<br>59.1) | -190 | 6 | 226 | 7 | 220 | 6 d to 6 d |
| Lopinavir/ritonavir-TOGETHER<sup>18</sup> | 4.8 | 5.7 | -0.9 (-<br>4.9, 3.1) | -18.4 (-155.4,<br>45.1) | -112 | 11 | 227 | 14 | 244 | NO difference by<br>Cox proportional HR |
| Tenofovir Disproxil<br>Fumarate Plus<br>Emtricitabine-AR0-CORONA<sup>50</sup> | 3.3 | 6.7 | -3.3 (-<br>14.3, 7.7) | -100 (-<br>1989.8, 80.9) | -30 | 1 | 30 | 2 | 30 |  |
| Metformin-COVID-OUT<sup>16</sup> | 3.2 | 1.3 | 1.8 (0.1,<br>3.5) | 57.5 (3.8,<br>81.3) | 55 | 19 | 601 | 8 | 596 | NO difference |
| Metformin - TOGETHER<sup>51</sup> | 11.8 | 11.2 | 0.7 (-5.5, 6.8) | 5.6 (-60.8, 44.6) | 152 | 24 | 203 | 24 | 215 | not reported |
| Fluvoxamine-TOGETHER<sup>21</sup> | 12.8 | 10.1 | 2.7 (-0.5, 5.9) | 21.1 (-4.8, 40.6) | 37 | 97 | 756 | 75 | 741 | NO difference- 40% resolved by day 14 |
| Fluvoxamine-STOP COVID<sup>52</sup> | 8.3 | 0.0 | 8.3 (1.9, 14.7) | 1 (1, 1) | 12 | 6 | 72 | 0 | 80 | YES (100% vs 91.7% resolved on day 7) p=0.009 |
| Fluvoxamine-COVID-OUT<sup>16</sup> | 1.7 | 2.0 | -0.3 (-2.5, 1.9) | -17.6 (-281, 63.7) | -333 | 5 | 293 | 6 | 299 | No difference (14 symptoms on 4 pt scale over 14 days) |
| Fluvoxamine ACTIV-6<sup>53</sup> | 0.3 | 0.1 | 0.18 (-0.4, 0.7) | 54.7 (-398.3, 95.9) | 555 | 2 | 607 | 1 | 670 | 12 d to 13 d |
| Fluvoxamine/budesonide-TOGETHER<sup>54</sup> | 1.1 | 0.9 | 0.1 (-0.9, 1.1) | 12.5 (-140.1, 68.1) | 738 | 8 | 738 | 7 | 738 | not reported |
| Ivermectin-TOGETHER<sup>55</sup> | 14.0 | 11.6 | 2.4 (-1.2, 5.9) | 16.8 (-9.9, 37.1) | 42 | 95 | 679 | 79 | 679 | NO difference- 40% resolved by day 14 |
| Ivermectin-COVID-OUT<sup>16</sup> | 1.4 | 1.1 | 0.3 (-1.3, 1.9) | 23.9 (-181, 79.4) | 299 | 5 | 356 | 4 | 374 | No difference (14 symptoms on 4 pt scale over 14 days) |
| Ivermectin Iran<sup>56</sup> | 5.0 | 7.1 | -2.1 (-6.1, 1.9) | -42.3 (-178, 27.2) | -47 | 14 | 281 | 19 | 268 | NO difference |
| Ivermectin-ACTIV-6<sup>57</sup> | 1.2 | 1.2 | -0.1 (-1.1, 1.0) | -5.3 (-158, 57.0) | -1634 | 9 | 774 | 10 | 817 | No difference (12d vs 13 d) |
| Ivermectin high dose-ACTIV-6<sup>58</sup> | 0.3 | 0.8 | -0.5 (-1.4, 0.4) | -150.1 (-1188, 51.1) | -200 | 2 | 604 | 5 | 602 | 11 d vs 11 d no difference |
| Hydroxychloroquine-TOGETHER<sup>18</sup> | 4.8 | 3.7 | 1.1 (-2.7, 4.9) | 22.9 (-88.1, 68.4) | 90 | 11 | 227 | 8 | 214 | NO difference by Cox proportional HR |
| Hydroxychloroquine-COVID-19 PEP<sup>59</sup> | 4.7 | 2.4 | 2.4 (-1.1, 5.9) | 50.2 (-43.1, 82.7) | 42 | 10 | 211 | 5 | 212 | NO Difference in symptom severity score over 14 days |
| Hydroxychloroquine-AH COVID-19<sup>60</sup> | 0.0 | 3.6 | -3.6 (-7.1, -0.1) | NA | -28 | 0 | 37 | 4 | 111 | NO difference 14 d vs 12 d |
| Hydroxychloroquin<br>e-BCN PEP-CoV-<br>2<sup>61</sup> | 7.0 | 5.9 | 1.1 (-4.5,<br>6.7) | 16.0 (-103,<br>65.2) | 89 | 11 | 157 | 8 | 136 | NO difference 10 d<br>vs 12 d |
| Hydroxychloroquin<br>e-BMG<sup>62</sup> | 4.8 | 3.4 | 1.4 (-4.0,<br>6.9) | 29.9 (-154,<br>80.6) | 69 | 4 | 83 | 5 | 148 | NO difference 11 d<br>vs 12 d |
| Hydroxychloroquin<br>e-Utah<sup>63</sup> | 2.6 | 4.6 | -2.0 (-<br>6.2, 2.2) | -73.8 (-481.6,<br>48.0) | -51 | 4 | 151 | 7 | 152 | 6 d to 6 d |
| Hydroxychloroquin<br>e/Azithromycin-Brazil<sup>64</sup> | 2.4 | 2.4 | 0 (-6.5,<br>6.5) | 0 (-1447,<br>93.5) | 100 | 1 | 42 | 1 | 42 |  |
| Nitazoxanide-Romark<sup>22</sup> | 2.6 | 0.5 | 2.0 (-0.4,<br>4.5) | 78.8 (-79.7,<br>97.5) | 49 | 5 | 195 | 1 | 184 | Yes mild illness (13<br>d vs 18 d , p=0.01),<br>NO difference for<br>moderate illness |
| Colchicine-COLCORONA<sup>20</sup> | 5.8 | 4.7 | 1.2 (-0.1,<br>2.5) | 20.0 (-2.8,<br>37.7) | 86 | 131 | 2253 | 104 | 2235 | Not reported |
| Losartan-MN<sup>65</sup> | 1.7 | 5.2 | -3.5 (-<br>10.1, 3.1) | -205 (-2749,<br>67.3) | -29 | 1 | 59 | 3 | 58 |  |
| Niclosamide<sup>66</sup> | 2.9 | 0.0 | 2.9 (-2.7,<br>8.6) | 1 (1, 1) | 34 | 1 | 34 | 0 | 33 | NO difference 12 d<br>vs 15 d |
| Aspirin-ACTIV-4B<sup>67</sup> | 0.7 | 0.7 | 0.04 (-<br>1.9, 2.0) | 5.6 (-1395,<br>94) | 2448 | 1 | 136 | 1 | 144 | Not reported |
| 2.5-mg apixaban-ACTIV-4B<sup>67</sup> | 0.7 | 0.7 | -0.01 (-<br>2.0, 2.0) | -0.7 (-1494,<br>93.6) | -18360 | 1 | 136 | 1 | 135 | Not reported |
| 5-mg apixaban<br>ACTIV-4B<sup>67</sup> | 0.7 | 1.4 | -0.7 (-<br>3.1, 1.7) | -90.2 (-1974,<br>82.6) | -151 | 1 | 136 | 2 | 143 | Not reported |
| Sulodexide<sup>68</sup> | 29.4 | 17.7 | 11.7 (1.1,<br>22.3) | 39.7 (3.5,<br>62.3) | 9 | 35 | 119 | 22 | 124 | Not reported |
| Enoxaparin-ETHIC<sup>69</sup> | 10.5 | 11.4 | -0.9 (-<br>9.2, 7.4) | -8.6 (-131,<br>49.0) | -111 | 12 | 114 | 12 | 105 | Not reported |
| Enoxaparin-OVID<sup>70</sup> | 3.4 | 3.4 | -0.1 (-<br>3.3, 3.2) | -1.7 (-166,<br>61.2) | -1740 | 8 | 238 | 8 | 234 | Not reported |
| Inhaled Ciclesonide-COVIS<sup>71</sup> | 3.4 | 1.7 | 1.8 (-1.4,<br>4.9) | 51.4 (-85.2,<br>87.2) | 56 | 7 | 203 | 3 | 179 | 19 d to 19 d |
| Inhaled ciclesonide-COVERAGE<sup>72</sup> | 11.2 | 12.7 | -1.5 (-10.1, 7.1) | -13.5 (-134, 45.0) | -66 | 12 | 107 | 14 | 110 | NO difference 13 d vs 12 d |
| Zinc<sup>73</sup> | 6.0 | 8.6 | -2.6 (-12.4, 7.2) | -43.7 (-471.4, 63.9) | -38 | 3 | 50 | 5 | 58 | 11 d to 10 d |
| Ascorbic acid<sup>73</sup> | 6.0 | 4.2 | 1.8 (-6.8, 10.5) | 30.6 (-297.6, 87.9) | 55 | 3 | 50 | 2 | 48 | 12 d to 10 d |
| Zinc/Ascorbic acid<sup>73</sup> | 6.0 | 12.1 | -6.1 (-16.7, 4.6) | -101.1 (-637, 45.1) | -16 | 3 | 50 | 7 | 58 | 10 d to 10 d |
| Homeopathy-COVID-Simile<sup>74</sup> | 6.8 | 2.4 | 4.4 (-4.3, 13.2) | 65.1 (-222.6, 96.2) | 23 | 3 | 44 | 1 | 42 |  |
| Saliravira<sup>75</sup> | 28.6 | 0.0 | 28.6 (16.7, 40.4) | 1 (1, 1) | 4 | 16 | 56 | 0 | 87 | YES 9d vs 14 d p<0.05 |
| Azithromycin-Atomic<sup>76</sup> | 11.6 | 10.3 | 1.2 (-5.9, 8.4) | 10.5 (-72.3, 53.6) | 82 | 17 | 147 | 15 | 145 | Not reported |
| Azithromycin-ACTION<sup>28</sup> | 0.0 | 4.0 | -4.0 (-7.4, -0.6) | NA | -25 | 0 | 72 | 5 | 125 | No difference resolution day 14 |
| Resveratrol<sup>77</sup> | 6.0 | 2.0 | 4.0 (-3.6, 11.6) | 66.7 (-210, 96.4) | 25 | 3 | 50 | 1 | 50 | Not reported |

## TRIAL FINDINGS

Examining RCTs by agent class, statistically significant relative risk reductions in hospitalisation were found in two of five CCP RCTs, six of nine mAb RCTs, four of 17 small molecule antiviral RCTs, but just 2 of 39 repurposed drug RCTs (Appendix Table 2). Except for the bebtelovimab RCT (2 hospitalisations in each arm^17^), mAb RCTs reduced the risk of hospitalisation by 50-80% (average 75%). Two of the three small molecule antiviral drugs (remdesivir^15^ and nirmatrelvir/ritonavir^5^) showed very high levels of relative risk reduction – 87% and 88% respectively – but molnupiravir reduced risk of hospitalisation by only 30%^7^ (no reduction in the PANORAMIC RCT^25^). The lopinavir/ritonavir combination was associated with a non-significant increase in risk of hospitalisation^18^.

**Table 2.**
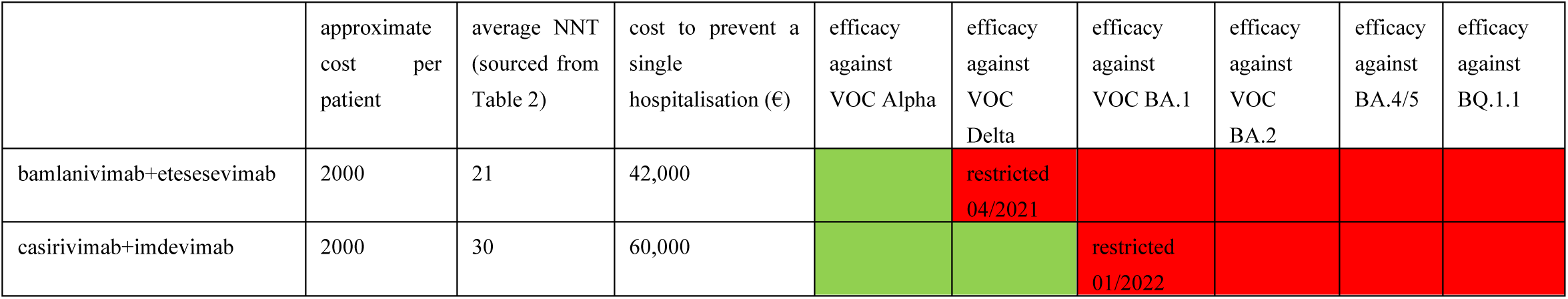
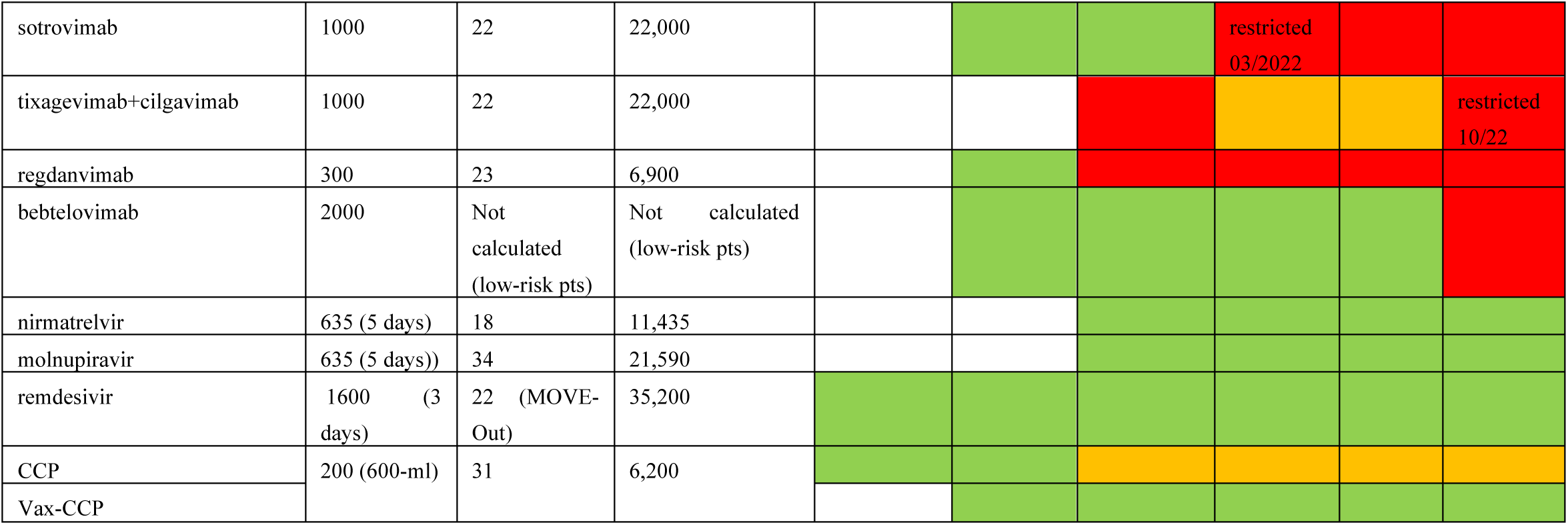
Summary of historical efficacy of different therapeutics against SARS-CoV-2 VOCs. White = drug not available at that time; green = effective; orange = partially effective; red= not effective. Restriction reported refer to initial restrictions by FDA. NNT: number needed to treat.

Among repurposed drug RCTs, all except metformin (58%) and sulodexide (40%), showed small and non-significant relative risk reductions of hospitalisation – ivermectin^19^, colchicine^20^, fluvoxamine^21^ and hydroxychloroquine^18^. The nitazoxanide^22^ RCT found one hospitalisation among 184 treated participants compared to five hospitalisations among 195 controls, too few events to achieve significance.

Absolute risk reductions (ARR) and number needed to treat (NNT) to avert hospitalisations varied across studies and treatment classes (Appendix Table 2). In general, except for the repurposed drugs the absolute risk difference approximated 3% if one excludes the molnupiravir-Panoramic Study from SMA. The CCP RCTs had an ARR of 3.2% (95%CI-0.9-5.6), mAbs RCTs had an ARR of 4.0% (95%CI-3.2-4.8), small molecule antivirals excluding molnupiravir-Panoramic had ARR of 2.1% (95%CI-1.3-2.9) and the repurposed drugs had a smaller ARR at 1.1% (95%CI-0.5-1.6). The number needed to treat to prevent hospitalisations approximated 30 in the trials, with a few notably low with CCP-Argentina (NNT=7) Nirmatrelvir/ritonavir (NNT=18), except those using repurposed drugs, where NNT averaged 70 (Appendix Table 2). In the pooled meta-analysis by class group, the CCP RCTs had a fixed effect OR of 0.69 (95% CI=0.53 to 0.9) with moderate heterogeneity (I^2^=43%), the mAbs had a fixed effect OR of 0.31 (95% CI=0.24-0.40) with low heterogeneity (I^2^=0%), the small molecule antivirals had a random effect OR of 0.78 (95% CI=0.48-1.33) with high heterogeneity (I^2^=69%) and the repurposed drugs had a random effect OR of 0.82 (95% CI-0.72-0.93) with low heterogeneity (I^2^=0) (Figure 4, Appendix Table 2). A biologic reason to use a fixed model for CCP and mAbs is that both formulations have the same active agent as specific antibody and thus there is a high degree of similarity in these two antibody interventions. Both antiviral small molecules and repurposed drugs comparison involve many types and classes of drugs for which the random effect model for the group is biologically more plausible than treating them as fixed. The meta-analysis of all interventions had a random effect OR of 0.67 (95% CI=0.57-0.80) with high heterogeneity (I^2^=52%) (Appendix Figure 2). Within the CCP RCTs excluding either CCP-Argentina for the nonhospital endpoint or CONV-ERT for methylene blue inactivation of antibody function only changed the OR by 0.05 with 95% CI remaining less than 1 for the fixed effect model (Appendix Figure 3A,B). Adding the 7 all-cause hospitalisations to CSSC-004, increased the OR by 0.02 and 95% CI by 0.01, essentially no change (Appendix Figure 3C).

**Figure 4.**
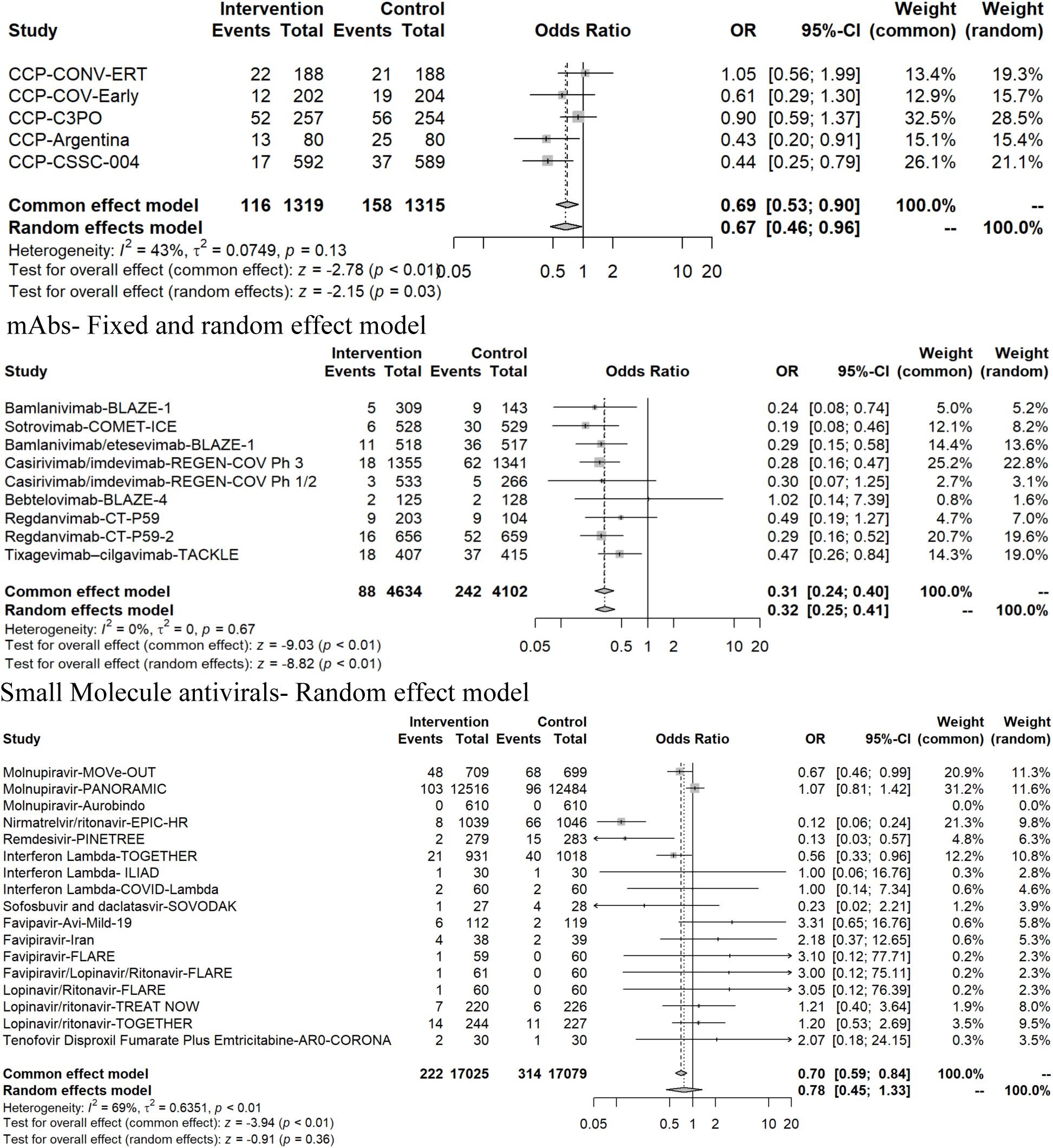
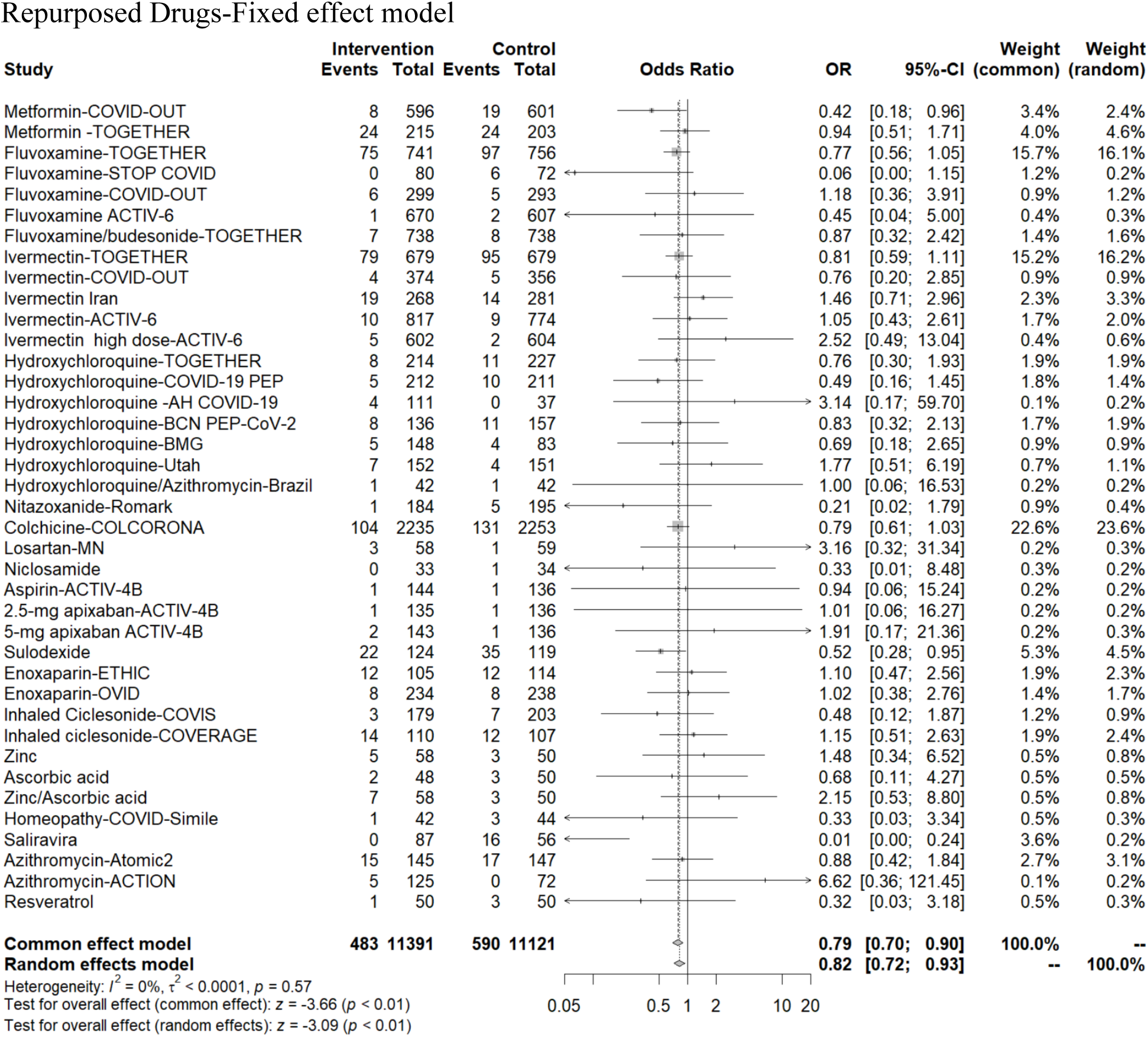
Odds ratio for hospitalisations with diverse therapeutic interventions, grouped according to mechanism of action (CCP, mAbs, small molecule antivirals and repurposed drugs).

Ten RCTs compared hospitalisation rates in early or late interventions (dated from symptom onset) that were extractable from the published papers or supplementary data as a post-hoc analysis (Figure 5). The small molecule drugs showed no significant reduction in the OR with segregation of early treatment from late treatment. In contrast, the antibody therapies showed a difference by treatment timing. Pooling the two classes of antibody treatment, the OR for early treatment was 0.65 (95%CI=0.49-0.85), while the OR for later treatment was 0.86 (95%CI=0.66-1.12). All four trial classes showed reduced rates of hospitalisation for each group. The final certainty of the available evidence with GRADE assessment (Appendix Table 4) showed a high certainty level within CCP trials, moderate certainty with mAbs, and low certainty with small molecule antivirals and repurposed drugs. The main reason for downgrading individual and pooled studies was imprecision, related to small number of participants and the wide confidence intervals around the effect, followed by ROB (Appendix Figure 4). We did not find concerns in any of the GRADE factors for CCP RCTs so we graded them as high level of certainty. mAbs were downgraded to moderate certainty due to ROB (in 4 of the 8 included RCTs, ROB for the outcome hospitalisation was judged of some concern). In the cumulative analysis, small molecule antivirals were downgraded to low certainty of evidence because of ROB (some/high ROB in 4 RCTs) and inconsistency (due to high heterogeneity), while repurposed drugs were downgraded to low certainty due to ROB (some/high ROB in 5 of the 11 comparisons) and indirectness (due to large difference in mechanism of action of the included drugs). The ROB was independently evaluated by the COVID-19-Network Meta-Analysis (NMA) initiative for most of the RCTs (Appendix Figure 4 and full output ROB in Supplementary Information). Funnel plot analysis shows a low risk of publication bias except for the mAbs, for which either the efficacy of high dose antibodies or non-reporting bias are plausible explanations (Appendix Figure 5).

**Figure 5.**
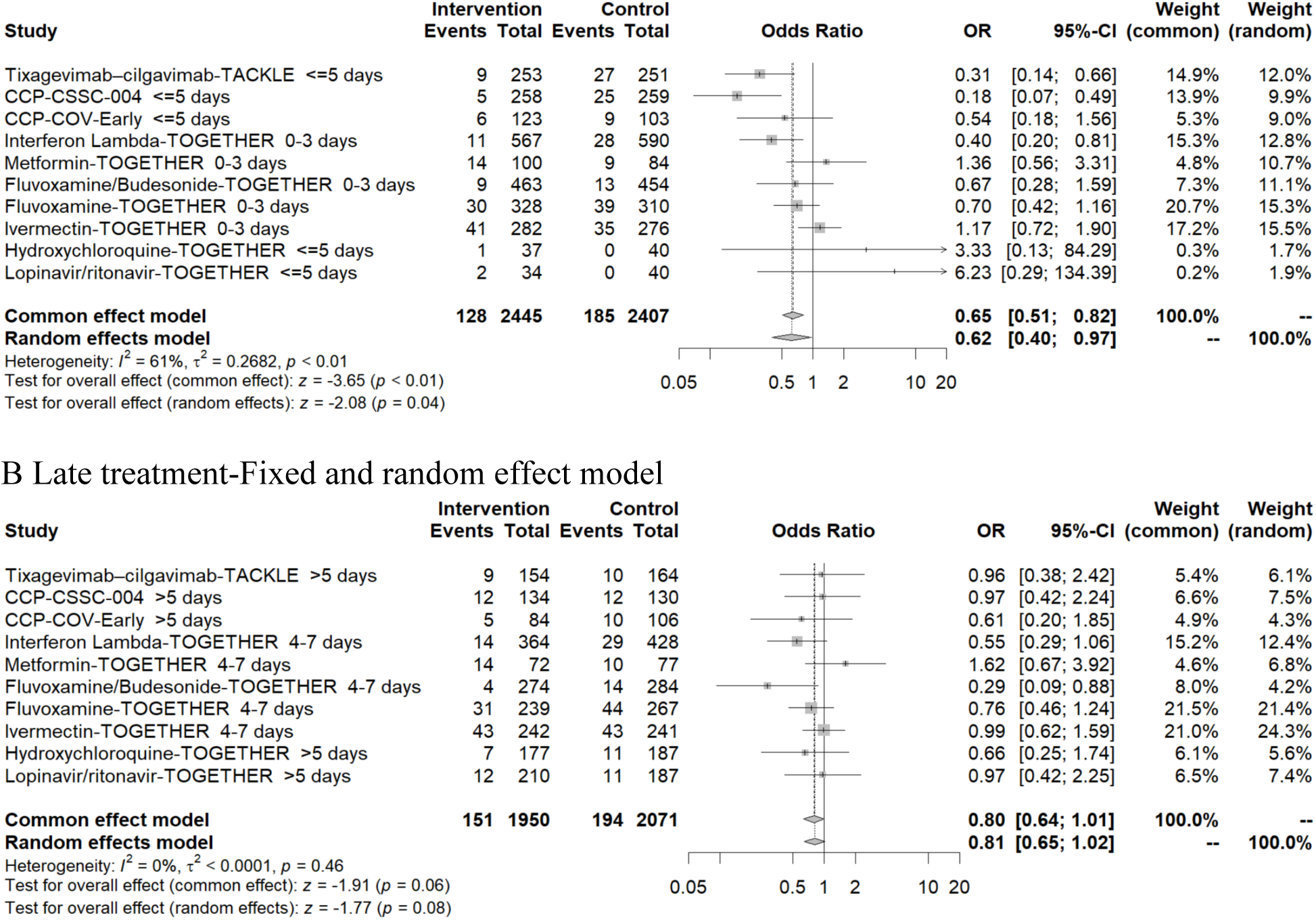
Odds ratio for point estimates for hospitalisation in RCT subgroups treated A) within 5 days since onset of symptoms and also B) over 5 days within same trial.

**Figure 6.**
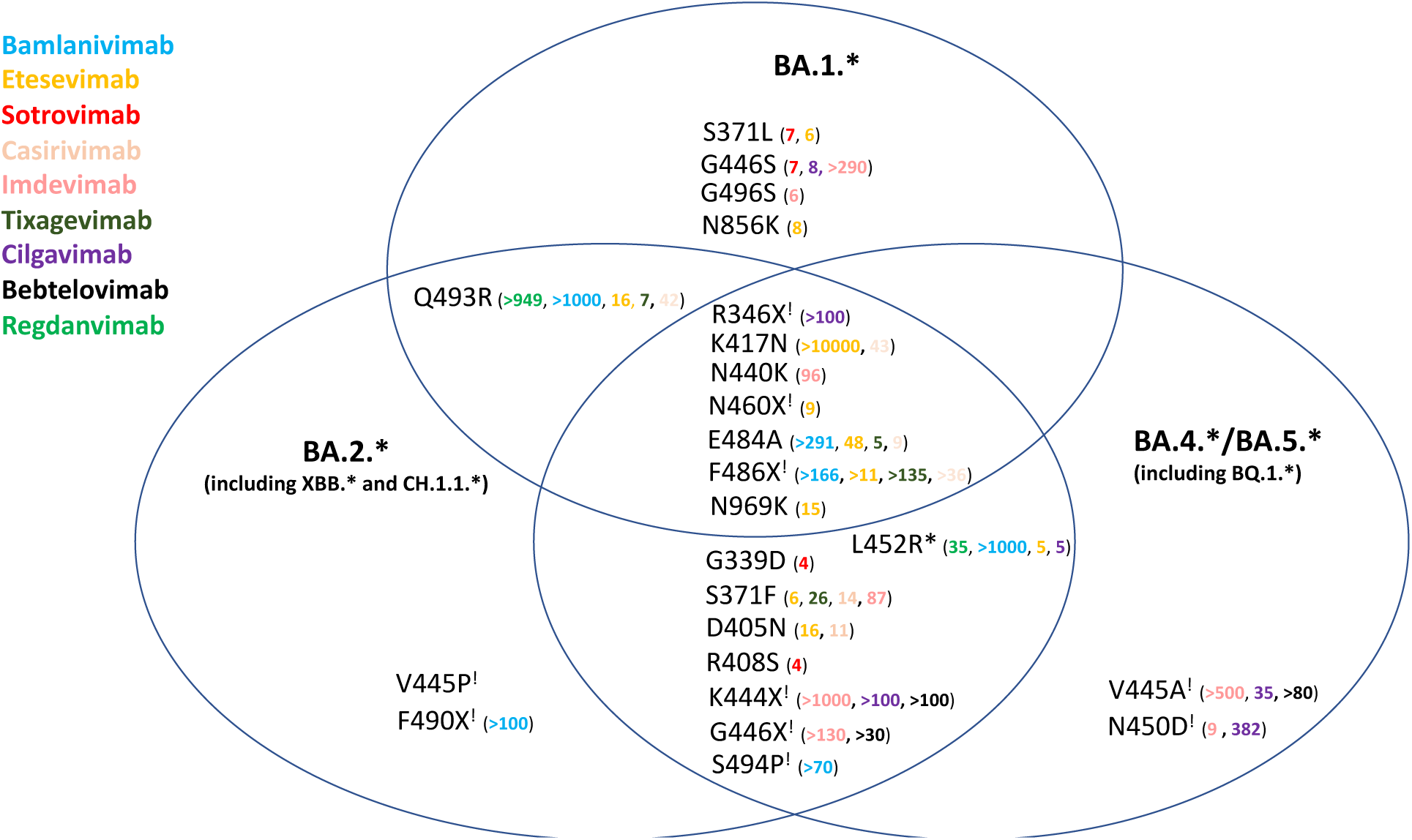
Venn diagram of mAb efficacy against Omicron sublineages. *In vitro* activity of currently approved mAbs against Omicron sublineages circulating as of October 2022. Specific Omicron Spike amino acid mutations causing baseline ≥ 4-fold-reduction in neutralization against mAbs are reported. Mutations for which the majority of studies are concordant are reported: the different fold-reductions for each mAb are identified across concordant studies as color coded numbers defining the mean median values of specific reduction in each study. Sourced from https://covdb.stanford.edu/page/susceptibility-data (accessed on January31, 2023* L452R occurs in all BA.4/BA.5 lineages, but only in several BA.2. sublineages.^! R346X^ and K444X occur in a growing number of BA.2 and BA.4/5 sublineages as a result of convergent evolution.

While several RCTs showed fewer deaths in the treatment arm, no outpatient study was powered to compare differences in mortality. Because of the low rate of deaths during trials the absolute risk reductions amongst the 4 antiviral classes are all below 1% corresponding to relative risk reductions of 20%, 81%, 87% and 22% for CCP, mAbs, small molecule antivirals or repurposed drugs, respectively (Appendix Table 4).

## TIME TO RESOLUTION

The two most effective CCP RCTs (Argentine^10^ and CSSC-004^9^) did not compare time to symptom resolution, while the COV-Early^23^ and ConV-ert^24^ RCTs reported no difference in the median time of symptom resolution in the two groups^24^ (Appendix Table 2). The mAbs noted faster resolution by 1, 2, 3, 4 or 5 days for bamlanivimab/etesevimab^25^, bebtelovimab^17^, regdanvimab^26^, casirivimab/imdevimab^2^, or redanvimab^27^ respectively. The smaller bamlanivimab-only RCT did not show a difference^1^. Of the three SMAs that noted reductions in hospitalisations, molnupiravir was associated with no difference in time of symptom resolution in MOVe-OUT^7^ but improvements in both PANORAMIC^25^ and Aurobindo^27^ RCTs. The 3-day outpatient remdesivir RCT showed that symptoms were alleviated by day 14 nearly twice as often^15^. The nirmatrelvir/ritonavir RCT did not report on this parameter^5^. Seven of 10 RCTs in the antiviral group did not show faster symptom resolution with intervention. The three RCTs largely performed in Brazil for fluvoxamine, ivermectin^19^ and hydroxychloroquine^18^ noted no differences in symptom resolution. Metformin did not evidence faster symptom resolution despite reducing hospitalisations. Three of the 25 RCTs reporting symptom resolution in the repurposed drug group noted faster symptom resolution.

## COSTS

mAbs and intravenous remdesivir schedules cost about 1000 to 2000 Euros per patient, respectively, while the oral drugs are much less than 1000 Euros per patient (Table 1). By comparison, the cost of CCP approximates 200 Euros per patient, and the cost for repurposed drugs is even lower. Considering the absolute risk reduction in hospitalisation, the number needed to treat to prevent a single hospitalisation is often very high, as are the associated costs. With the recently patented antivirals, costs for outpatient treatment often exceed the cost of a COVID-19 hospitalisation^28^.

mAbs successively lost efficacy against Delta and Omicron, with cilgavimab (the only Omicron-active ingredient in Evusheld™) and bebtelovimab also failing against BQ.1.1 and XBB.* sublineages (Figure 6). This has led the FDA to withdraw EUAs, while EMA has not restricted usage at all. Small molecule antivirals retain *in vitro* efficacy against Omicron, but concerns remain: molnupiravir showed low efficacy *in vivo*^6^ and is mutagenic for mammals *in vitro*^29^, while nirmatrelvir/ritonavir has drug/drug interaction contraindications (CYP3 metabolites especially tacrolimus, anti-cholesterol, anti-migraine or many anti-depressants) and has been associated with early virological and clinical rebounds in immunocompetent patients^30^. CCP from unvaccinated donors does not inhibit Omicron, but CCP from donors having any sequence of vaccination and recent, within 6 months, COVID-19 or having had boosted mRNA vaccine doses universally has high Omicron-neutralizing activity.

## DISCUSSION

The pharmaceutical industry – with well-established internal resources for trials and substantial economic support – was able to perform large outpatient trials of mAbs early in the pandemic. Inpatient services are generally more accessible to physician-scientists working in academic medical centers. The relative ease of conducting inpatient RCTs may have led most initial CCP, small molecule antiviral and repurposed trials – which were conducted principally by academic institutions – to be based in hospitals, often in patients treated too late for antiviral treatment to be expected to work, given that antiviral therapy must be given early in disease. The constrained resources available for clinical research by academic medicine during pandemic conditions further interfered with trial work, and several potentially valuable RCT’s with promising findings were terminated before they could provide definitive data. The findings of such trials are reported as null but often viewed as negative, notwithstanding trends towards effectiveness, and are rarely incorporated into clinical recommendations.

Consistent with the above, outpatient RCT data confirm that most antiviral/antimicrobial therapies are more effective when given before hospital admission. In examining the full assembly of these effective, yet molecularly disparate interventions, we note the consistent importance of early outpatient treatment for patients at risk of progression to hospitalisation^31^. Treatment within 5 days of illness onset was more effective than later treatment, as would be expected for an antiviral mechanism of action. An individual participant meta-analysis of CCP that investigated the effects of early compared to late treatment and of high compared to low dose antibody levels found that both early treatment and high levels of antibody combined to most effectively reduce risk of hospitalisations^32^.

The paucity of head-to-head RCT comparisons amongst outpatient COVID-19 therapies, however, makes the choice of therapy difficult. The outpatient RCTs reviewed here were conducted during different time-periods during the pandemic, thus targeting different variants, and enrolled participants with different vaccination statuses. Further heterogeneity was contributed by variation across the 54 RCTs, in participant age, medical risk factors and serological status These limitations need to be considered in our head-to-head meta-analysis of COVID-19 outpatient placebo controlled RCTs.

The choice, however, has been narrowed in recent months, and the clinical armamentarium has been reduced to small molecule antivirals, repurposed drugs and CCP, because single and double (“cocktail”) MAbs have lost effectiveness against new VOCs^33^. Both vaccine-elicited and disease-elicited antibodies are polyclonal, meaning that they include various isotypes that provide functional diversity and target numerous epitopes making variant escape much more difficult with CCP. Hence, polyclonal antibody preparations are much more resilient to the relentless evolution of variants. This is in marked contrast to mAbs, which target single epitopes of SARS-CoV-2. The exquisite mAb (and receptor binding domain) specificity renders mAbs susceptible to becoming ineffective with single amino acid changes. Plasma from individuals who have been both vaccinated and boosted is characterized by high amounts of neutralizing antibodies which can be effective against practically any existing VOC, including Omicron^34^ (so-called “heterologous immunity”, likely due to the well-known phenomenon of “epitope spreading”). Vaccine-boosted CCP has more than ten times the amount of total SARS-CoV-2 specific antibody and viral neutralizing activity compared to the pre-omicron CCP used in the effective outpatient CCP RCTs.

In addition to efficacy, other points to consider in an outpatient pandemic are tolerability, scalability and affordability. Repurposed drugs are generally well tolerated, widely available and relatively inexpensive, but, as we have shown, have limited efficacy. By contrast, small molecule antivirals are often plagued by contraindications and side effects, which make several classes of patients reliant on passive immunotherapies. Both small molecule antivirals and mAbs take time to develop and are unaffordable in low-and-middle income countries (LMIC). CCP is instead a tolerable, scalable, and affordable treatment and is usually provided in a single IV session, in contrast to Remdesivir, which requires a three-day intravenous course.

On Dec. 28, 2021 the FDA expanded the authorized emergency use of convalescent plasma with high titers of anti-SARS-CoV-2 antibodies “for the treatment of COVID-19 in patients with immunosuppressive disease or receiving immunosuppressive treatment, in either the outpatient or inpatient setting.” This EUA noted that CCP was safe and effective in immunocompetent individuals and allowed under the emergency measure of the pandemic, but expanded its use in immunosuppressed individuals to outpatient use, notwithstanding the availability of oral drugs and (at that time) two remaining effective mAb treatments for the new omicron variants of concerns

The published mAbs RCTs assembled here showed better efficacy than other outpatient interventions, yet are now clinically ineffective against BQ.1.* and XBB.* Omicron variants. CCP and small molecule antivirals have comparable levels of effectiveness, but the latter have many contraindications and side effects. Repurposed drugs are either ineffective or mildly effective. That leaves CCP, which is especially important in the immunosuppressed but, as the trials show, early treatment with high levels of antibody, has value in other populations as well. Our clinical recommendation from this review is to use CCP on an out-patient basis in regions with no other therapy available regardless of vaccination status for those at high risk of progression to hospitalisation.

## CONTRIBUTORS

DS wrote the first draft and extracted data verified by DF, and MF. DF curated Table 3 and 7 and revised the text. MC, JO, MF and DS performed statistical analyses. MC and DS performed GRADE assessment. AC, NP, MF and DH critically revised the manuscript. DS and DF directly accessed and verified the underlying data reported here. All authors read and agree with manuscript.

## DATA SHARING

Datasets used for this systematic review are publicly available in PubMed, medRxiv and bioRxiv.

## Data Availability

Datasets used for this systematic review are publicly available in PubMed, medRxiv and bioRxiv.

## Appendix for

### Description of trial participants

The median age of participants was about 50 years. The CCP group had a nonweighted trial average of median age equal to 58 years, while the anti-Spike mAbs, small molecule antivirals and repurposed drug groups younger average of median age was equal to 45 to 48 years. Most RCTs had more women than men, and 84% of all RCT 60,043 participants had Caucasian ethnicity (Table 1, Appendix Table 2).

The individual RCTs differed in the percentage of participants with risk factors for progression to severe COVID-19. Of the 37 RCTs reporting aggregated hospitalization risk factors, ten had 100% of participants with at least one hospitalization risk factor, while 5 had fewer than 50%. The bebtelovimab placebo-controlled RCT explicitly focused exclusively on low-risk individuals ^1^. Individual risk factors such as diabetes mellitus occurred in 10 to 20% of participants in most RCTs. Obesity with BMI over 29 averaged near 40% of RCT participants in the 4 therapy groups after excluding the large single 25,000 molnupiravir-PANORAMIC RCT with 15% of participants with BMI’s over 30 (Table 1) ^25^.

Of 18 RCTs reporting seropositivity rates at baseline, 11 had < 25% screening seropositive (Table 1, Figure 2). The molnupiravir-PANORAMIC RCT was an outlier, with 98% participant seropositives^25^. All but one^2^ of the RCTs enrolled within 8 days (median) of symptom onset. In RCTs of anti-Spike mAbs and small molecule antivirals, median time from illness onset to intervention was 3.5 to 4 days (Figure 3, Table 1, Appendix Table 2). CCP and repurposed antiviral drug RCTs enrolled within 4.5 to 5.1 days from symptom onset.

The CCP RCTs were conducted in the USA^3, 4^, Argentina^5^, Netherlands^6^ and Spain^7^ (Appendix Table 2). The anti-Spike mAb RCTs all had a USA component, but were largely centered in the Americas except for the sotrovimab RCT, which took place in Spain^8^. Many of the repurposed drugs and nirmatrelvir/ritonavir RCTs recruited worldwide^9^.

Four of the five CCP RCTs (COV-Early^6^, CONV-ERT^7^, Argentina^5^ and C3PO^4^), and all eight anti-Spike mAb RCTs took place in the setting of the D614G variant and the Alpha VOC (Figure 2). By contrast, most of the molnupiravir, nirmatrelvir/ritonavir^9^ and interferon lambda RCTs were conducted in the setting of the Delta VOC. The ivermectin^10^ and fluvoxamine^11^ RCTs ended as the Delta VOC wave began in August 2021. The remdesivir RCT spanned D614G, Alpha and Beta VOC but missed Delta^12^. The CSSC-004 RCT of CCP was the longest RCT reviewed, spanning periods characterized by D614G to Delta VOC infections^3^.

**Appendix Figure 1.**
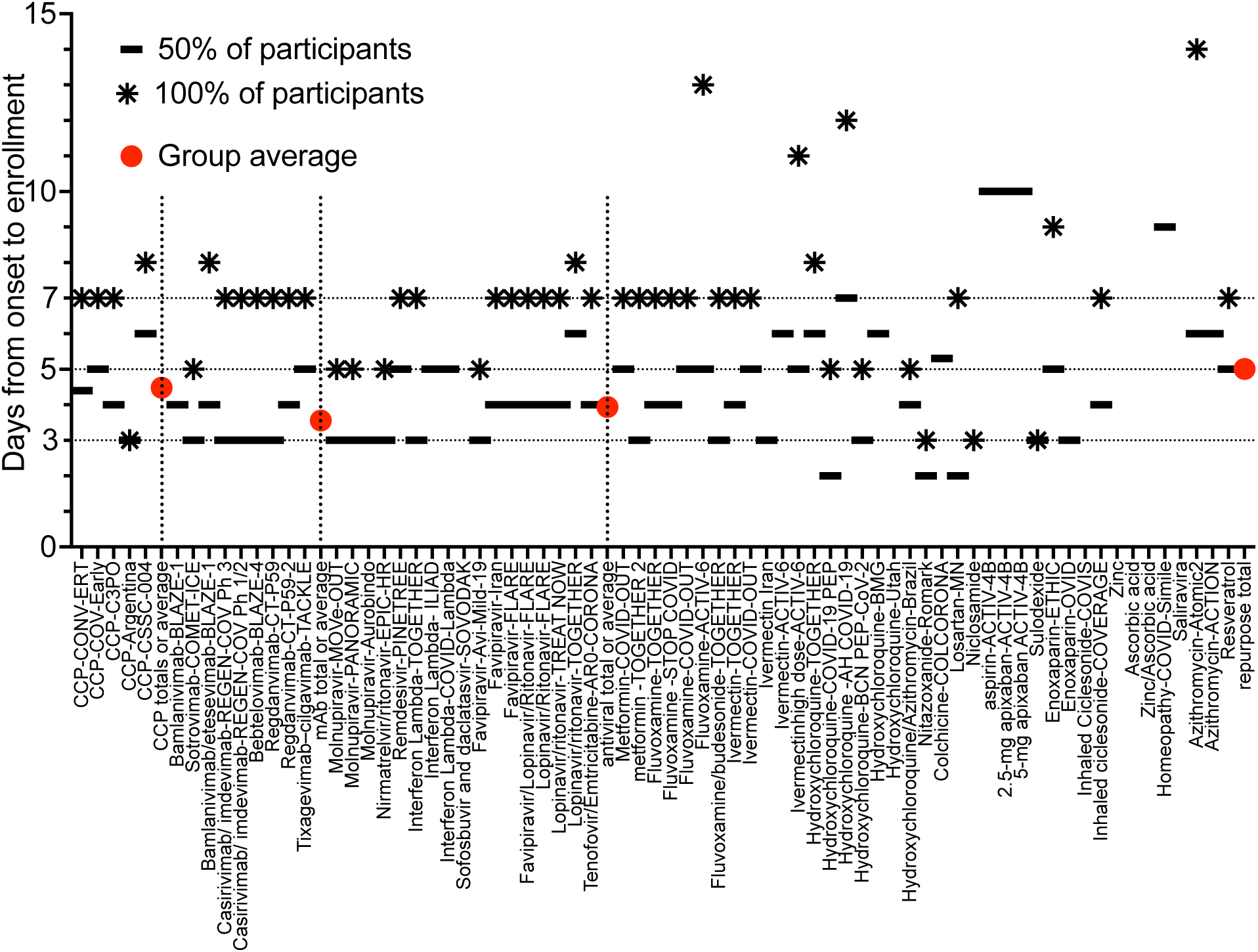
Comparison of mean interval from symptom onset to enrollment/intervention as well as per protocol interval inclusion limit for all participants.

**Appendix Figure 2:**
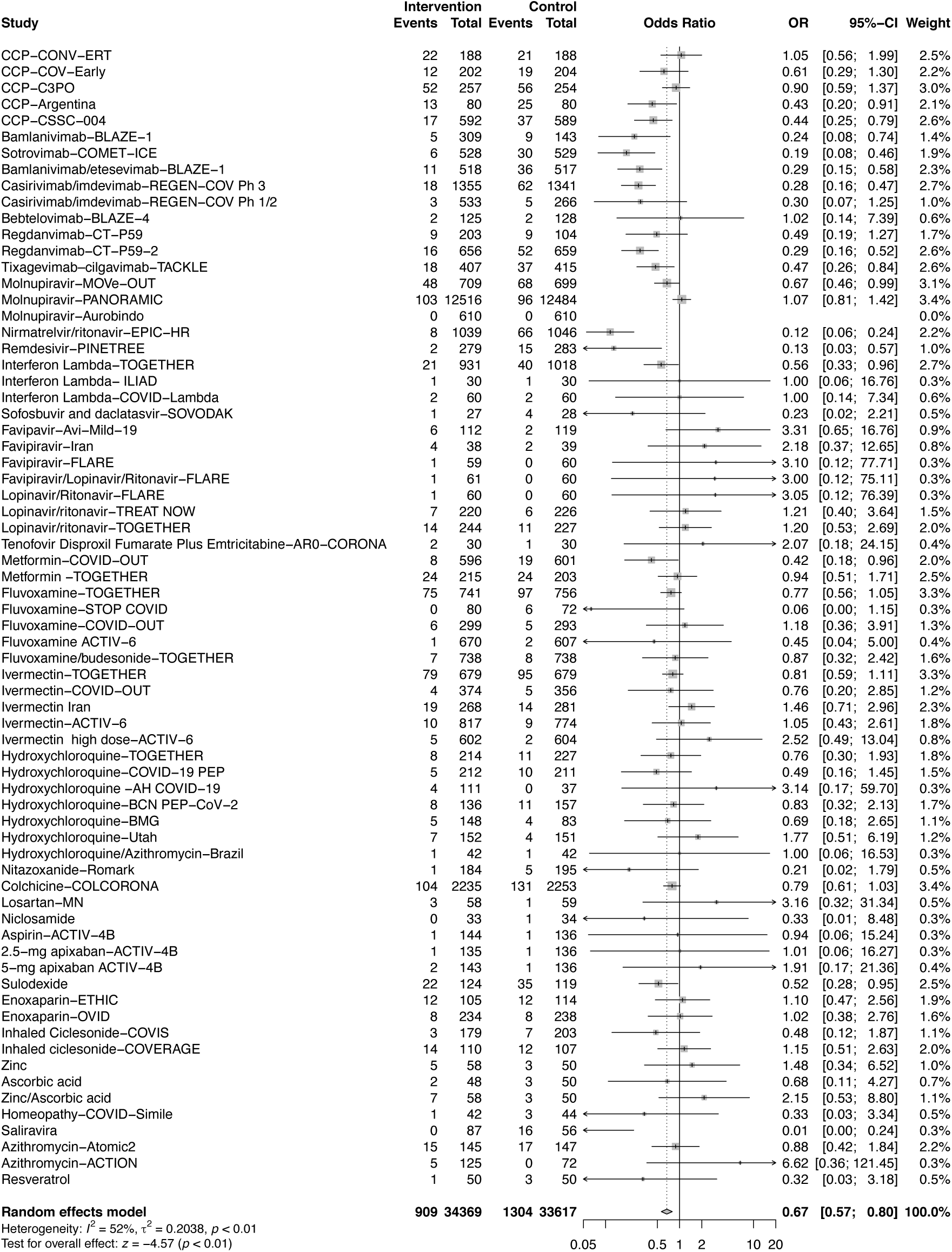
Odds ratio for hospitalizations from all interventions. Therapeutic interventions ordered according to mechanism of action (CCP, anti-Spike mAbs, small molecule antivirals and repurposed drugs

**Appendix Figure 3:**
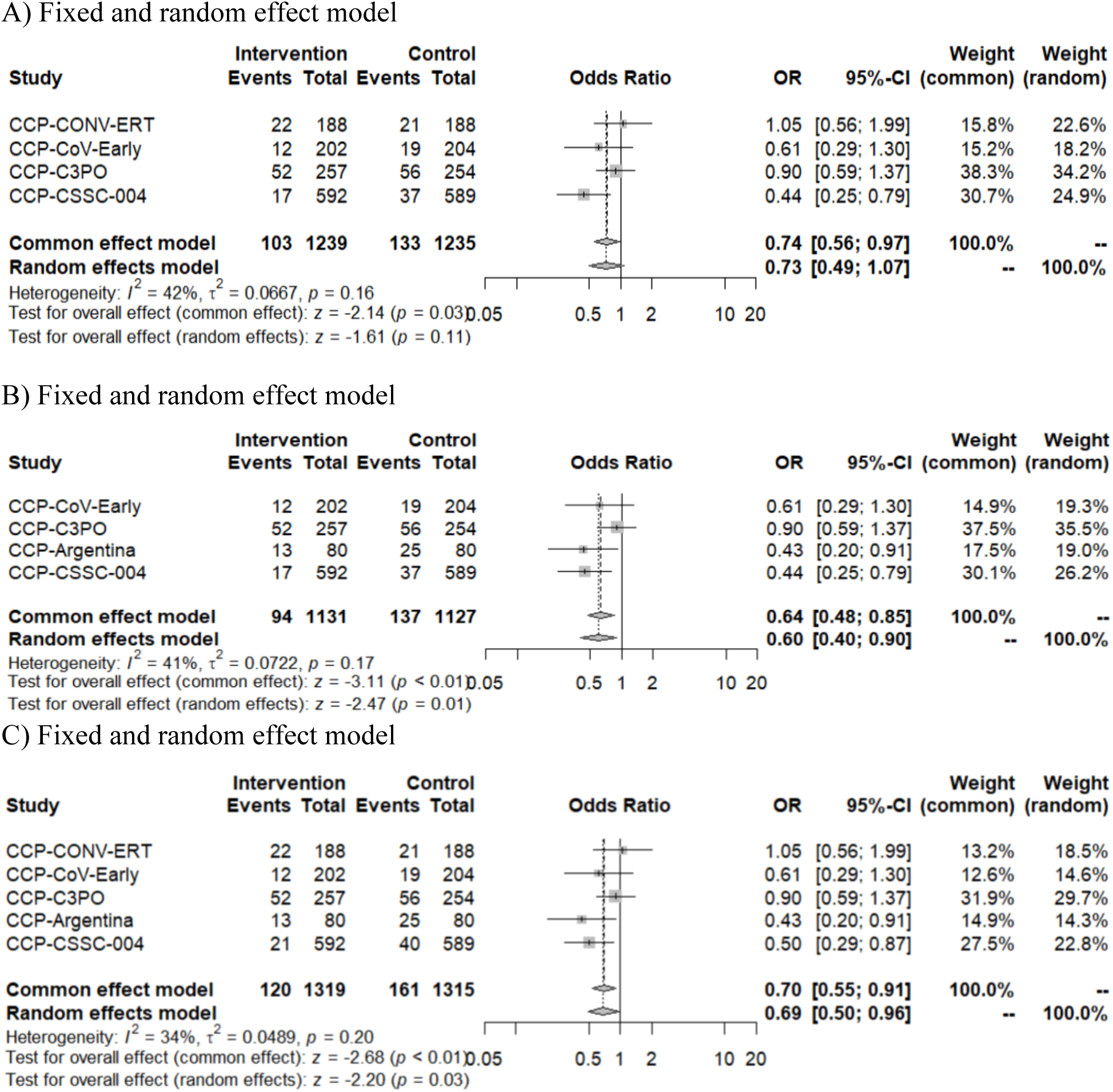
Odds ratio for hospitalizations within CCP group. A) All CCP trials excluding CCP-Argentina with a non hospital endpoint of severe respiratory distress or B) All CCP excluding CONV-ERT because of methylene blue inactivation of antibody function. C) all cause hospitalization for CSSC-004 to match all cause hospitalization for other CCP studies

**Appendix Figure 4:**
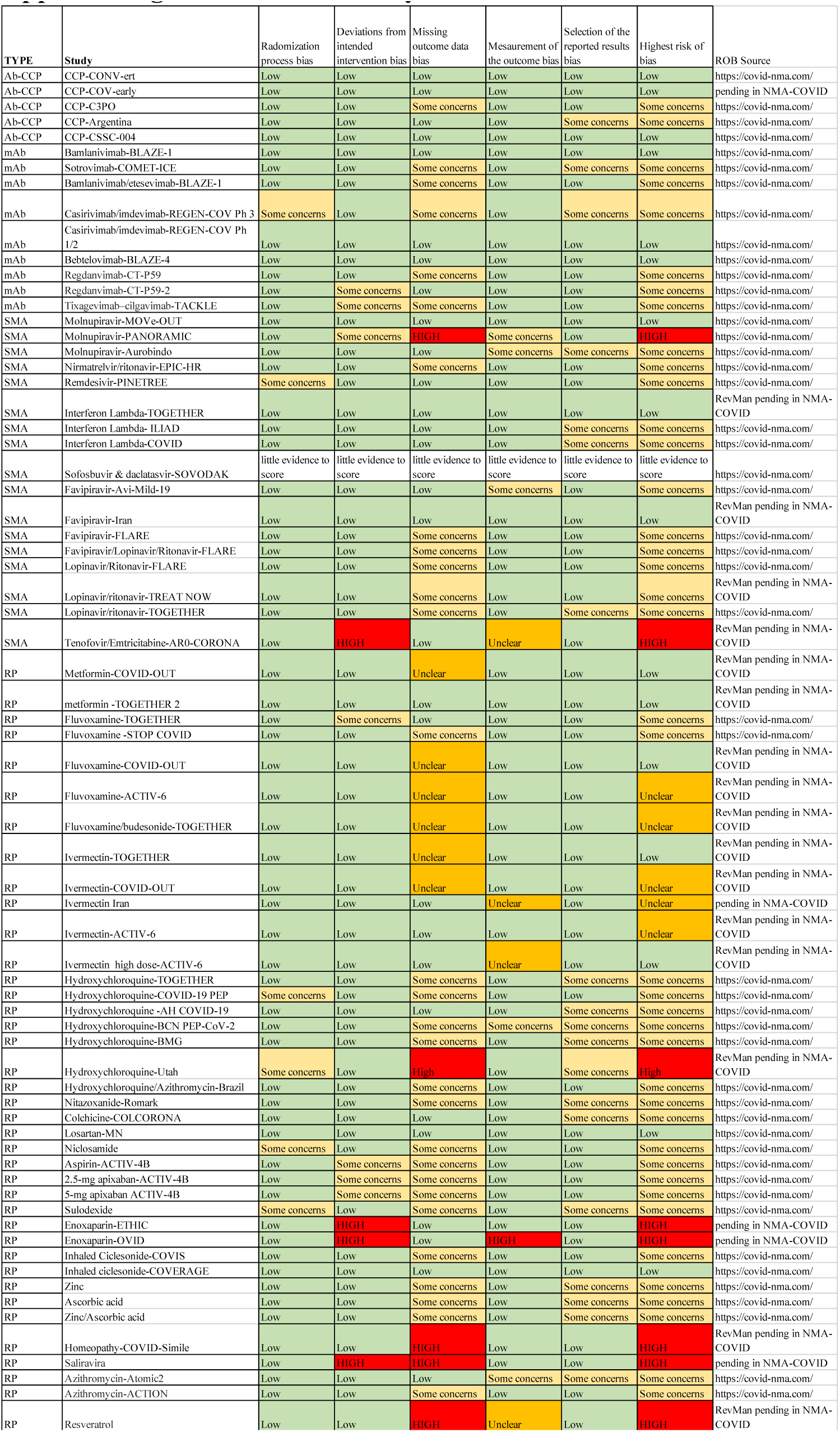
Risk of bias by RCT.

**Appendix Figure 5:**
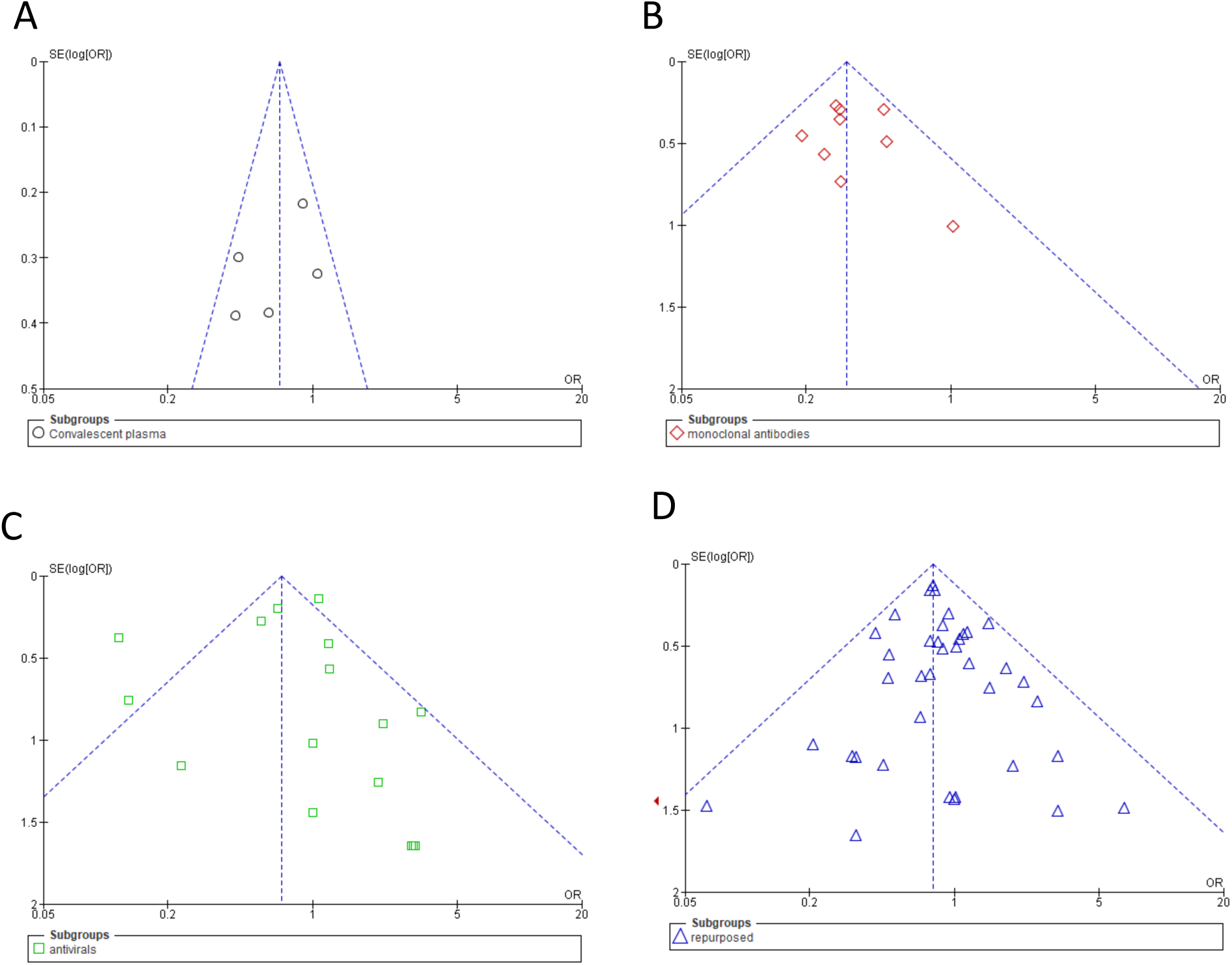
Funnel plots by RCTs class. A) CCP, B) anti-Spike mAbs C) small molecule antivirals and D) repurposed drugs. For anti-Spike mAbs RCTs, there is a suggestion of missing studies on the right side of the plot, where results would be unfavourable to the experimental intervention, for which either very high efficacy of high-dose anti-Spike mAbs or non-reporting bias is a plausible explanation.

**Table 1a.**
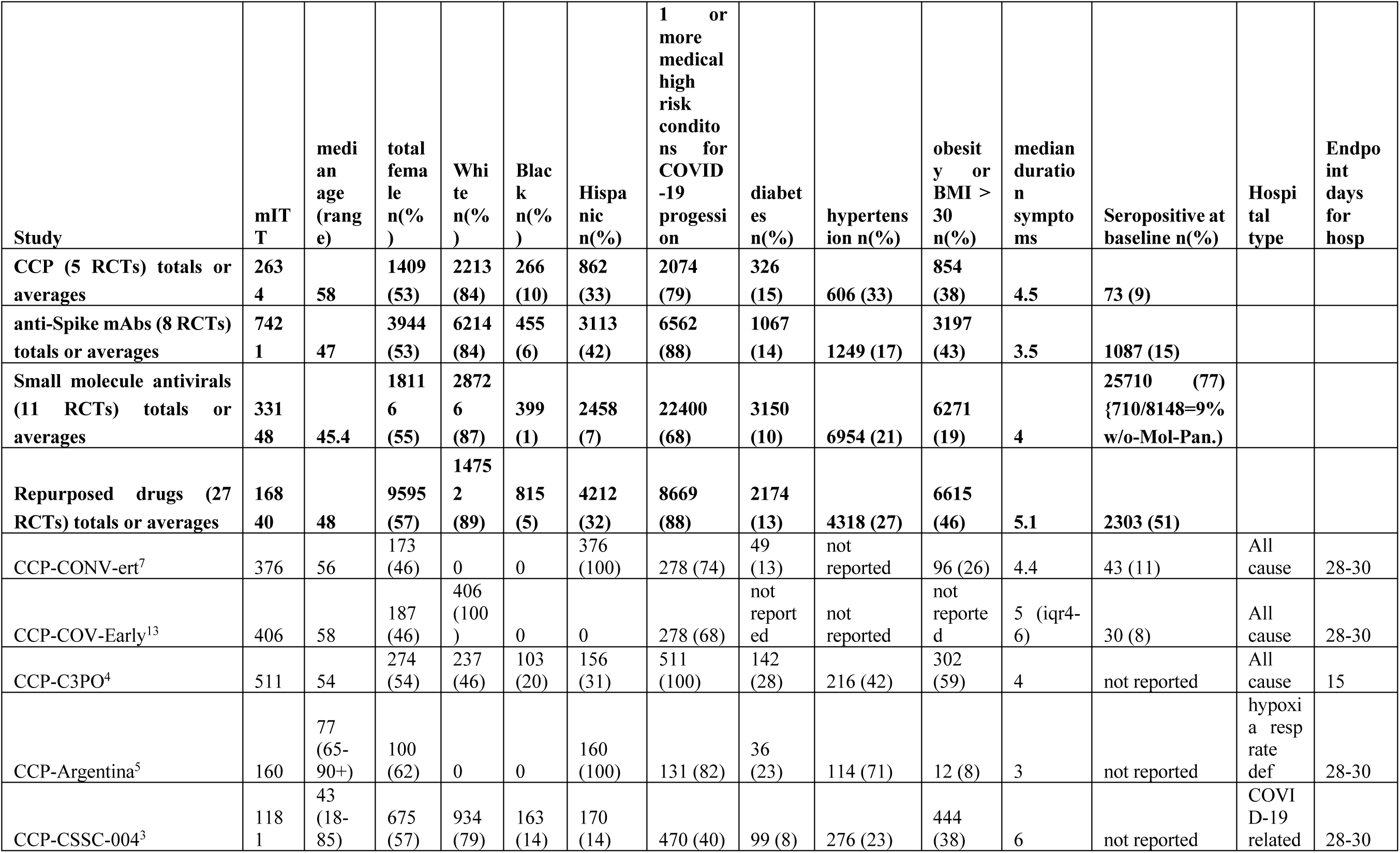
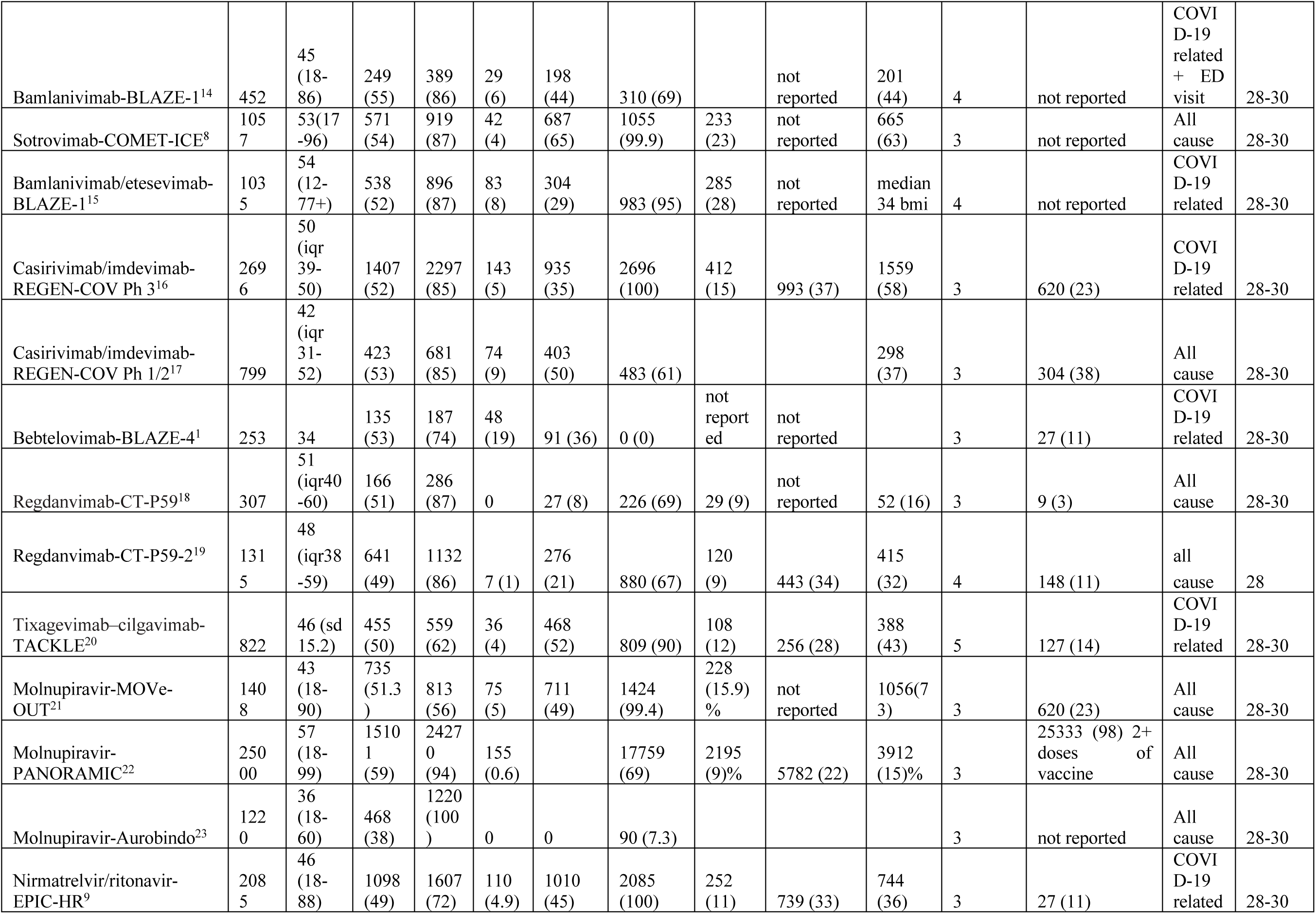
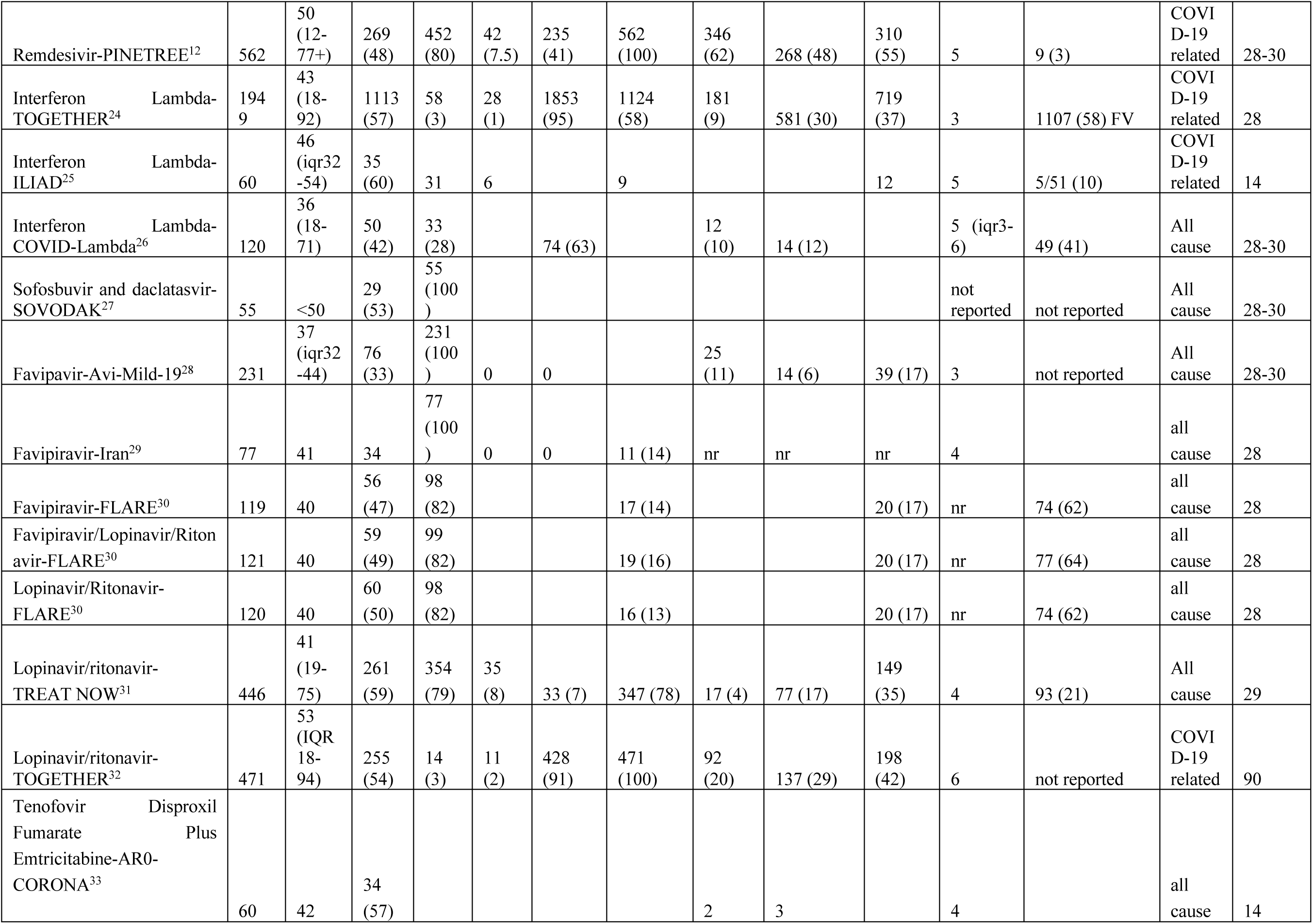
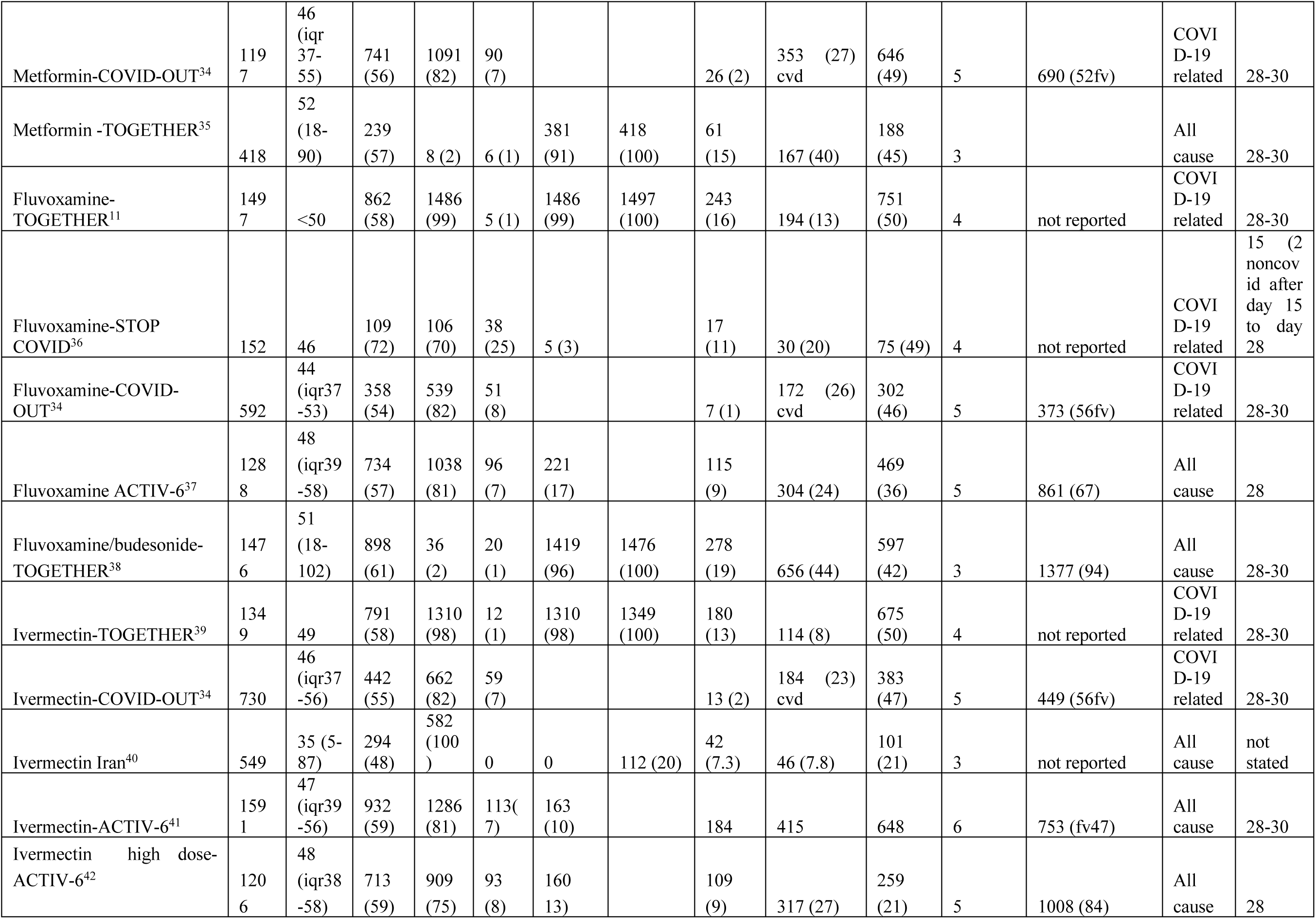
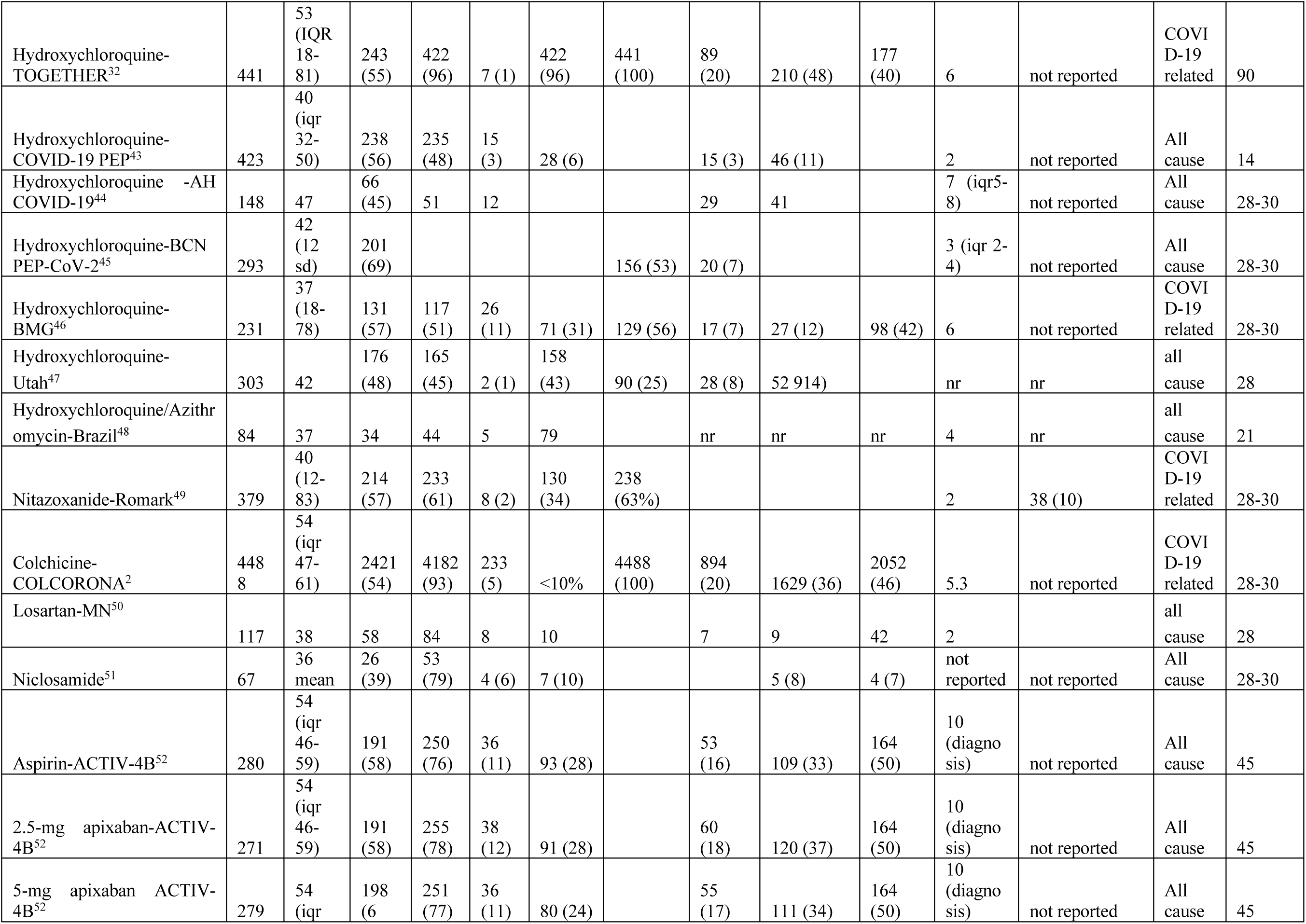
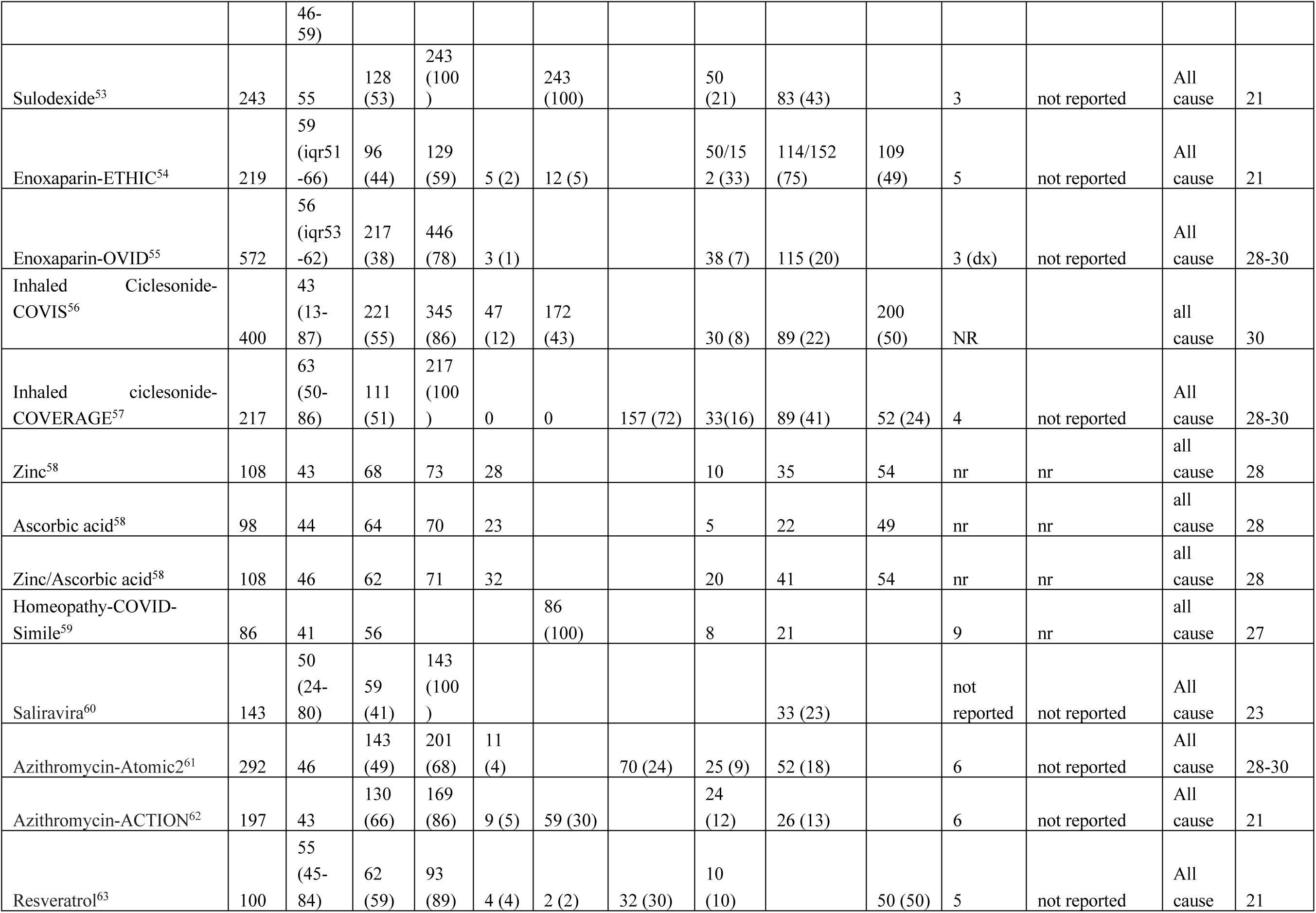
Demographic and clinical characteristics of recruits in the RCTs analyzed in this review.

**Appendix Table 1b:**
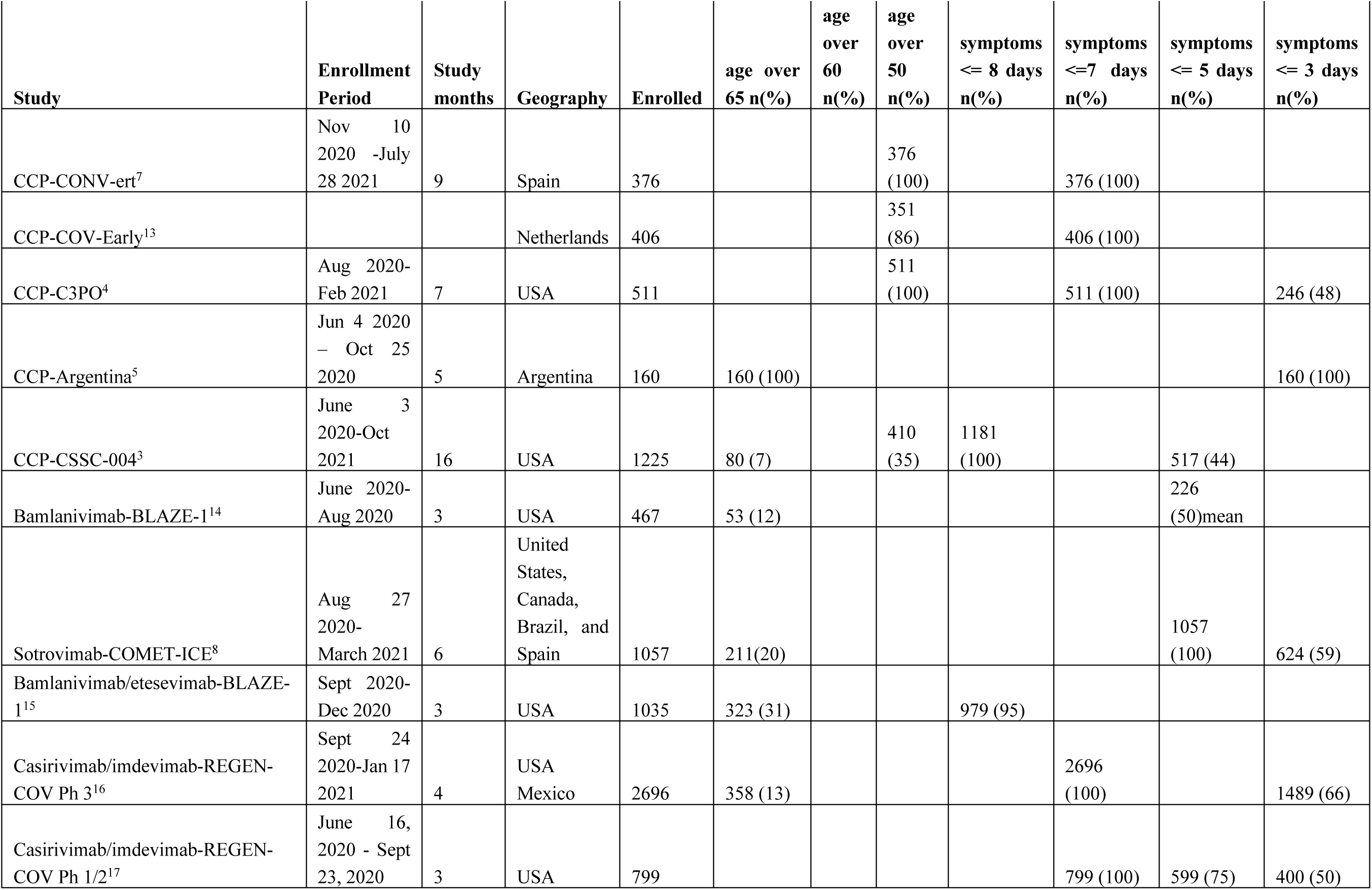
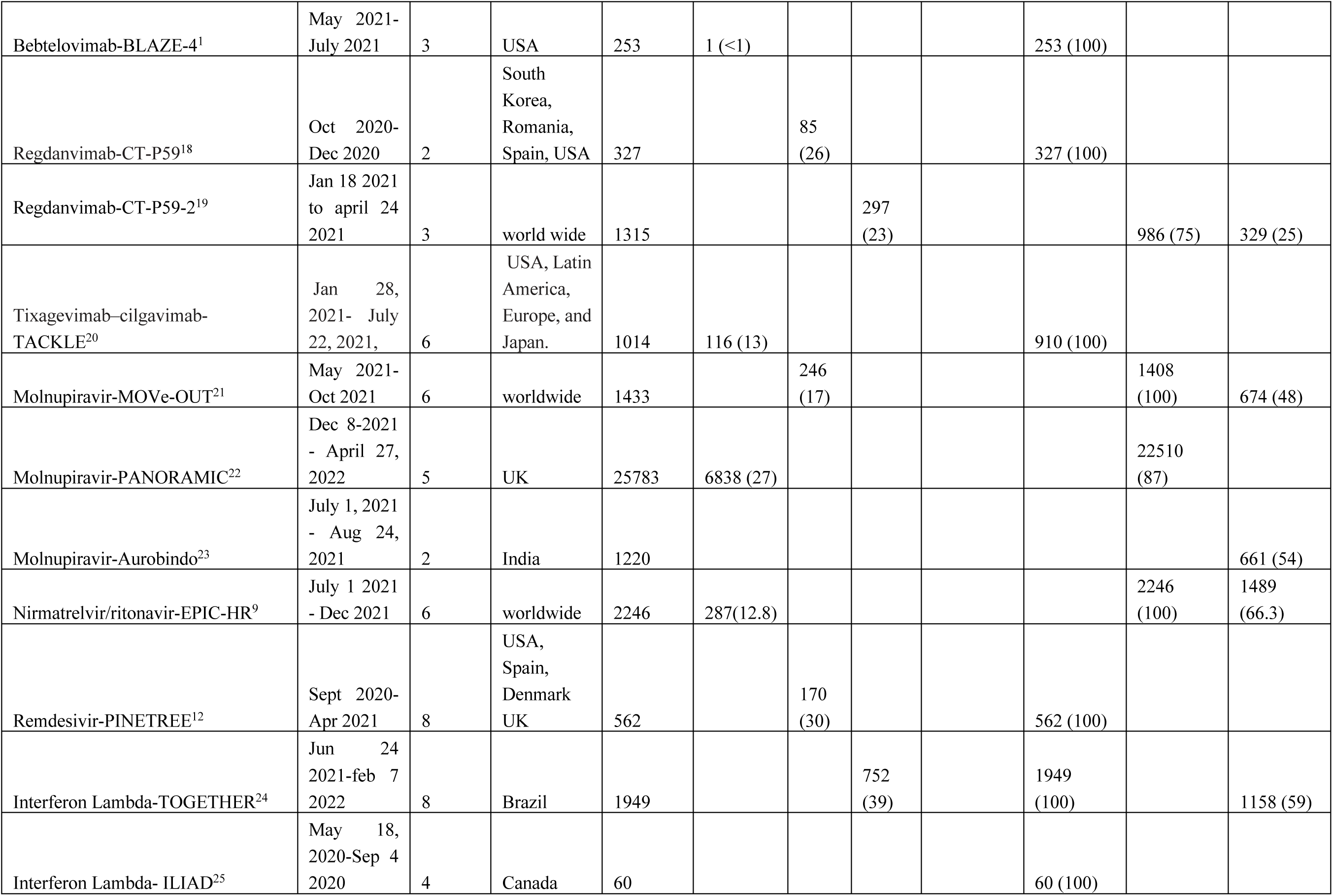
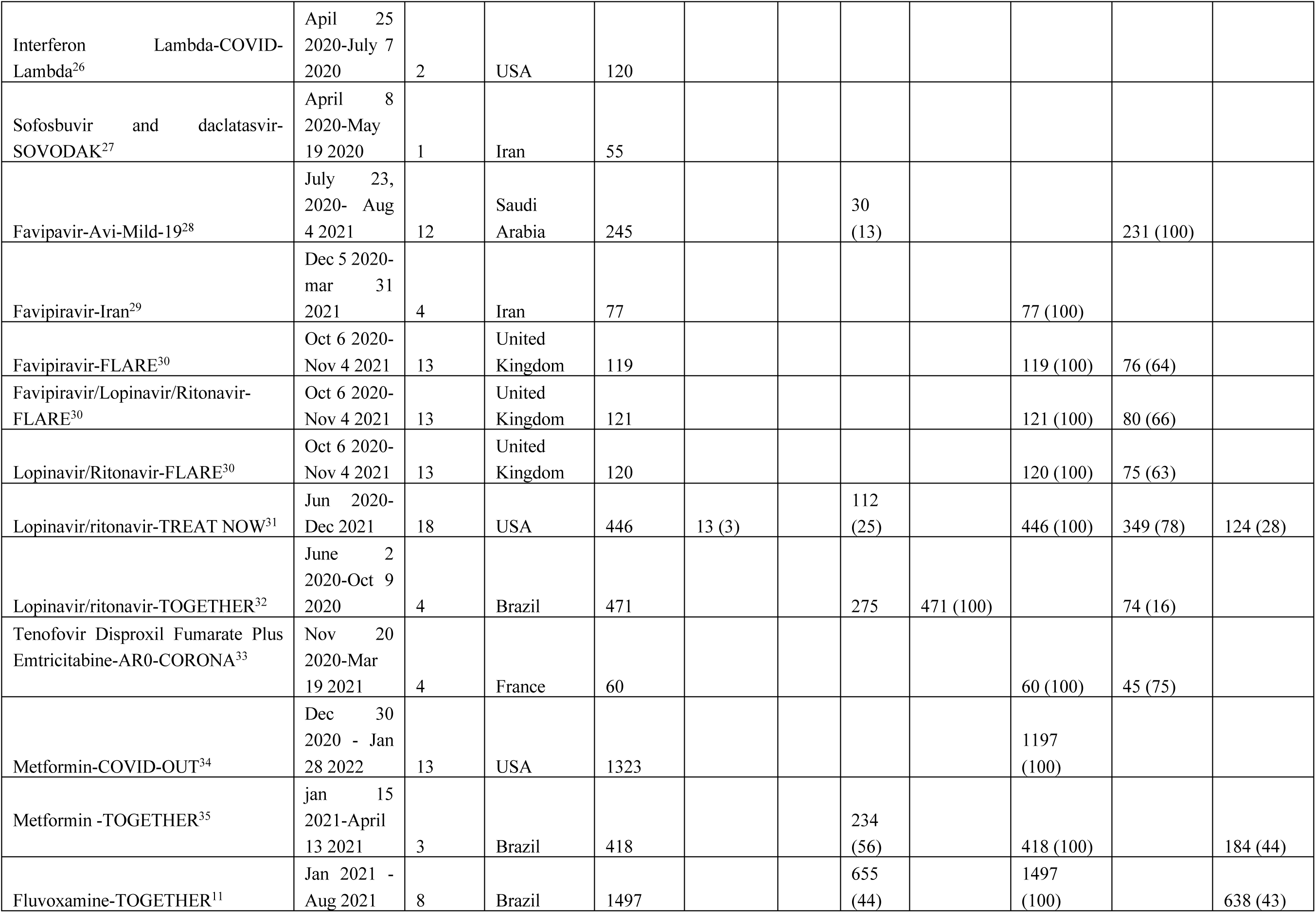
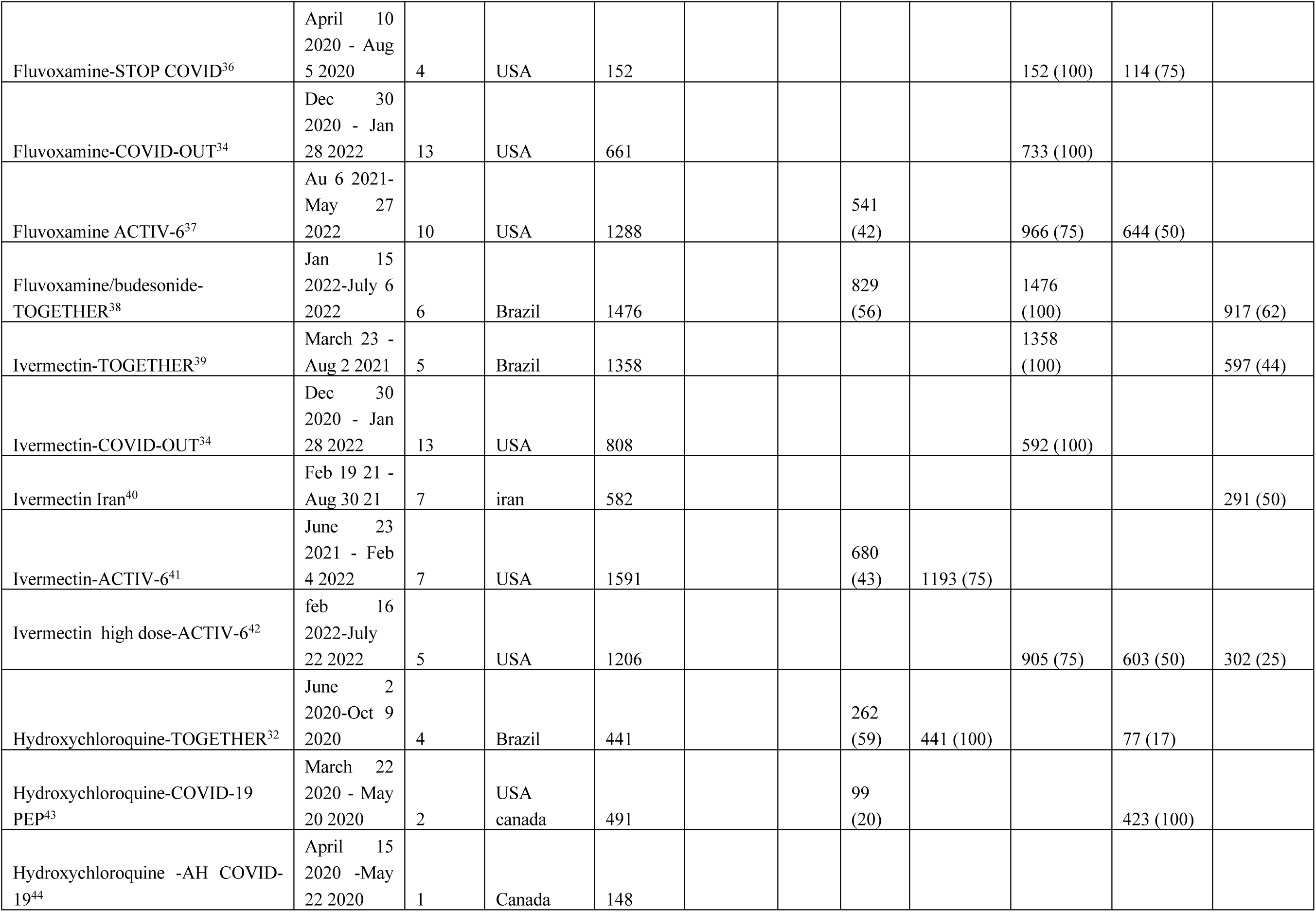
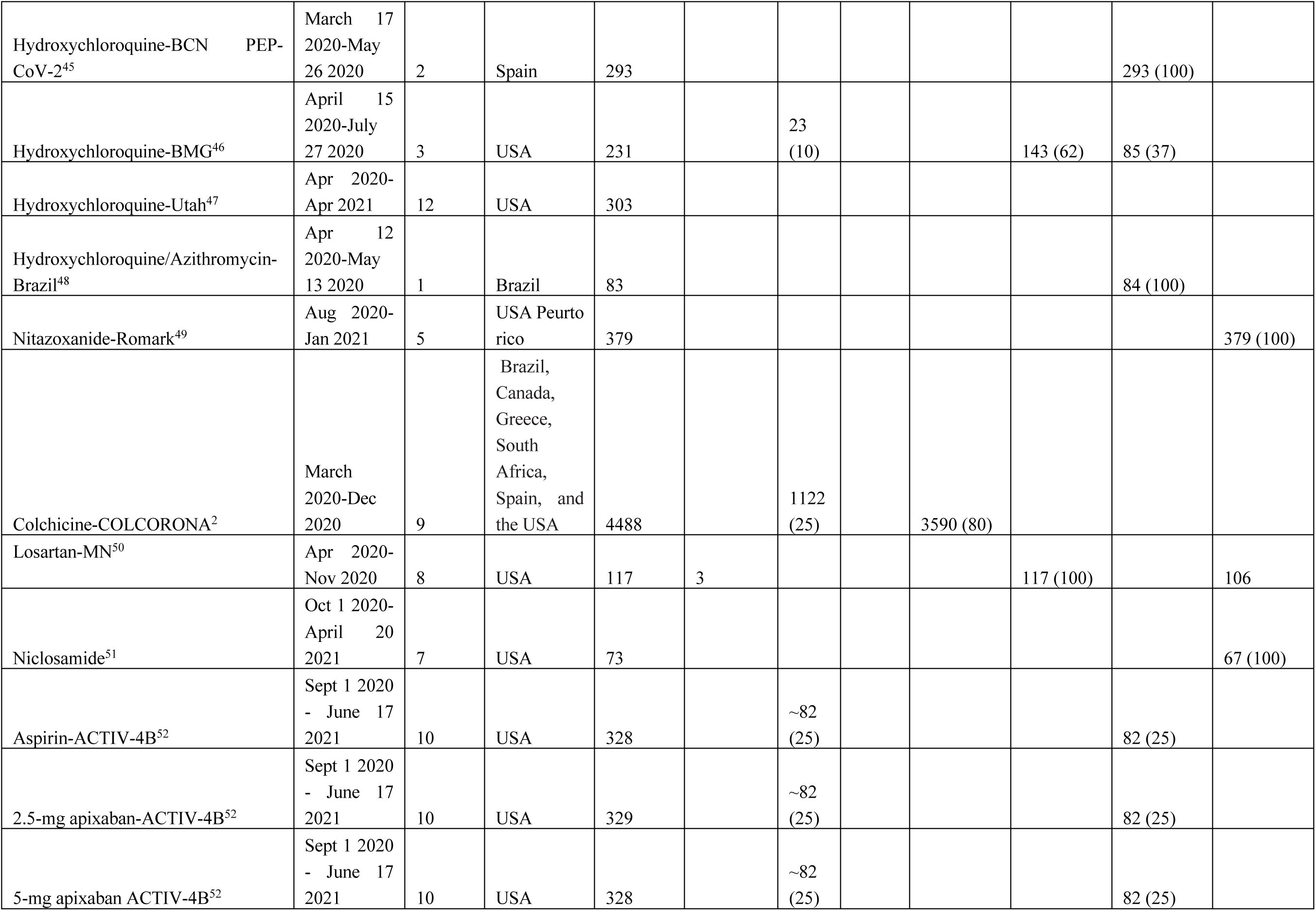
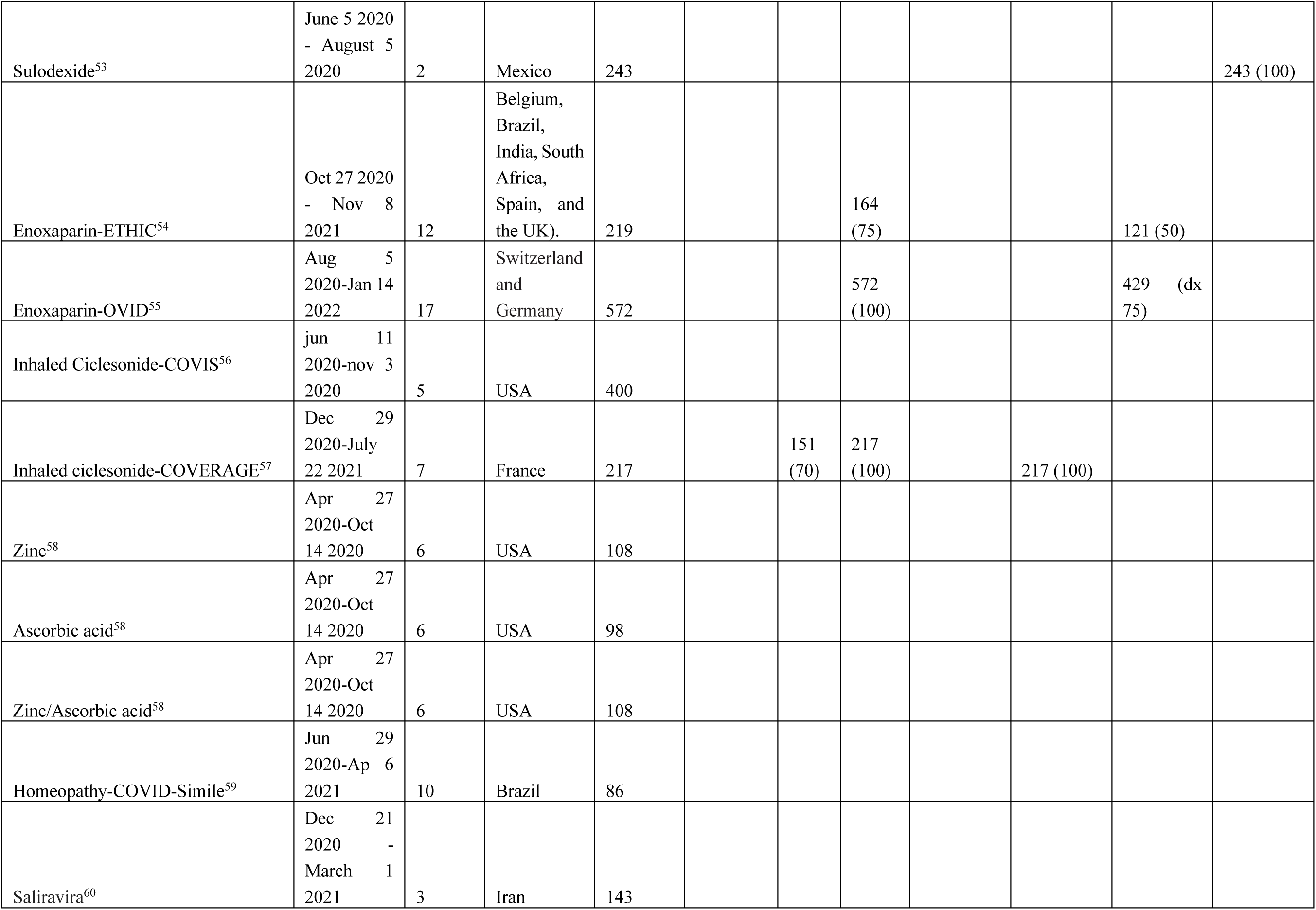
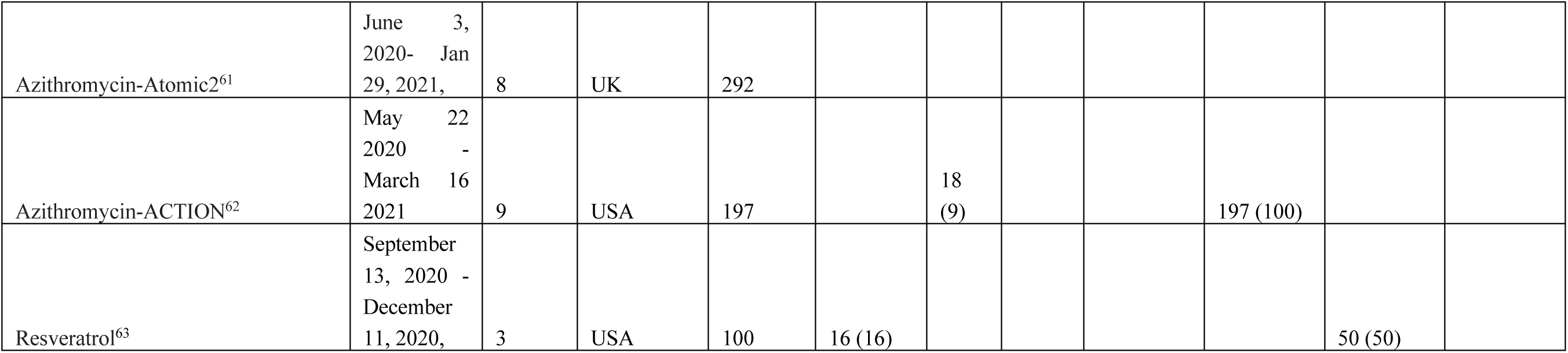
Additional baseline data from RCTs-Geography, age symptom onset.

**Appendix Table 2.**
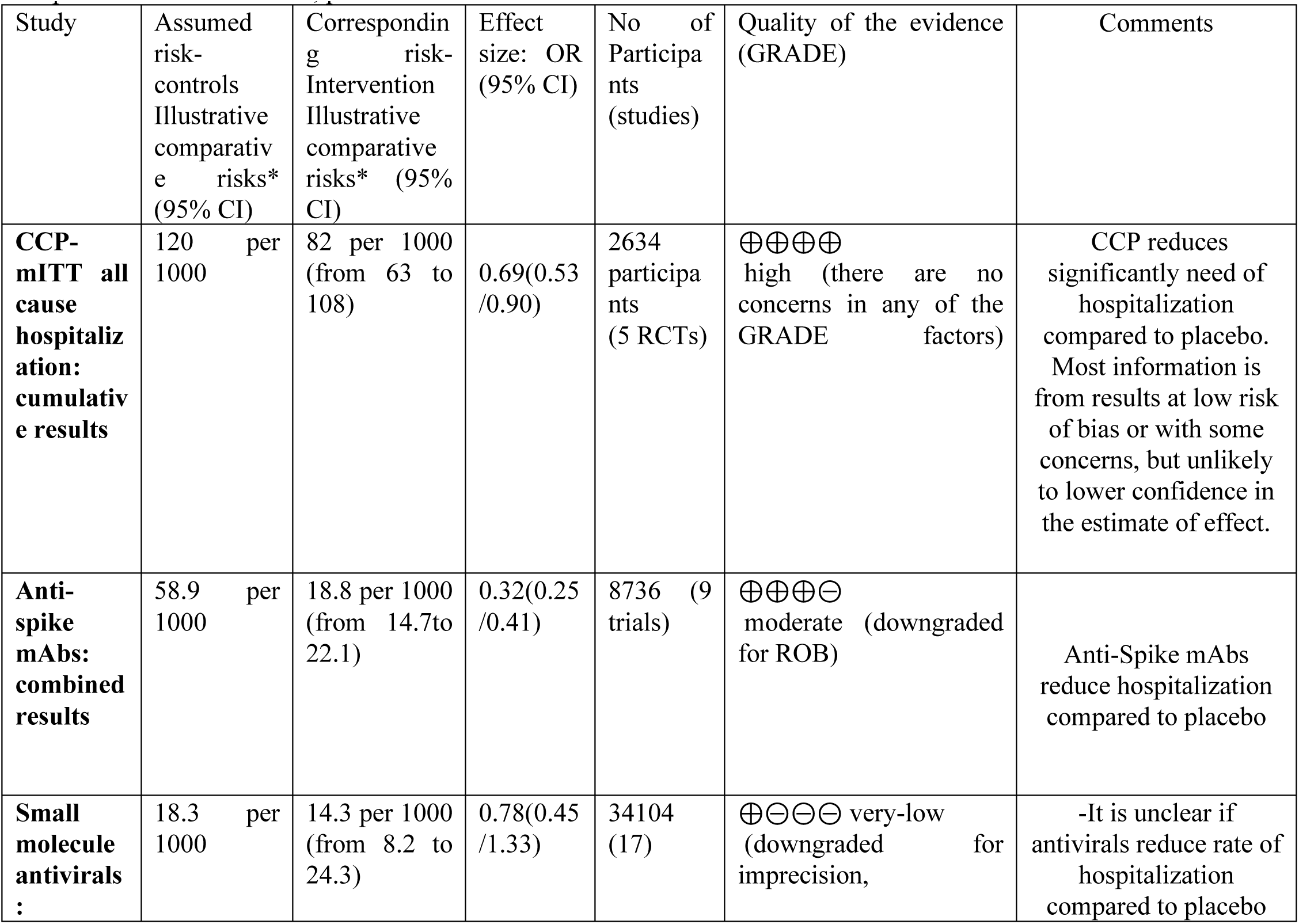
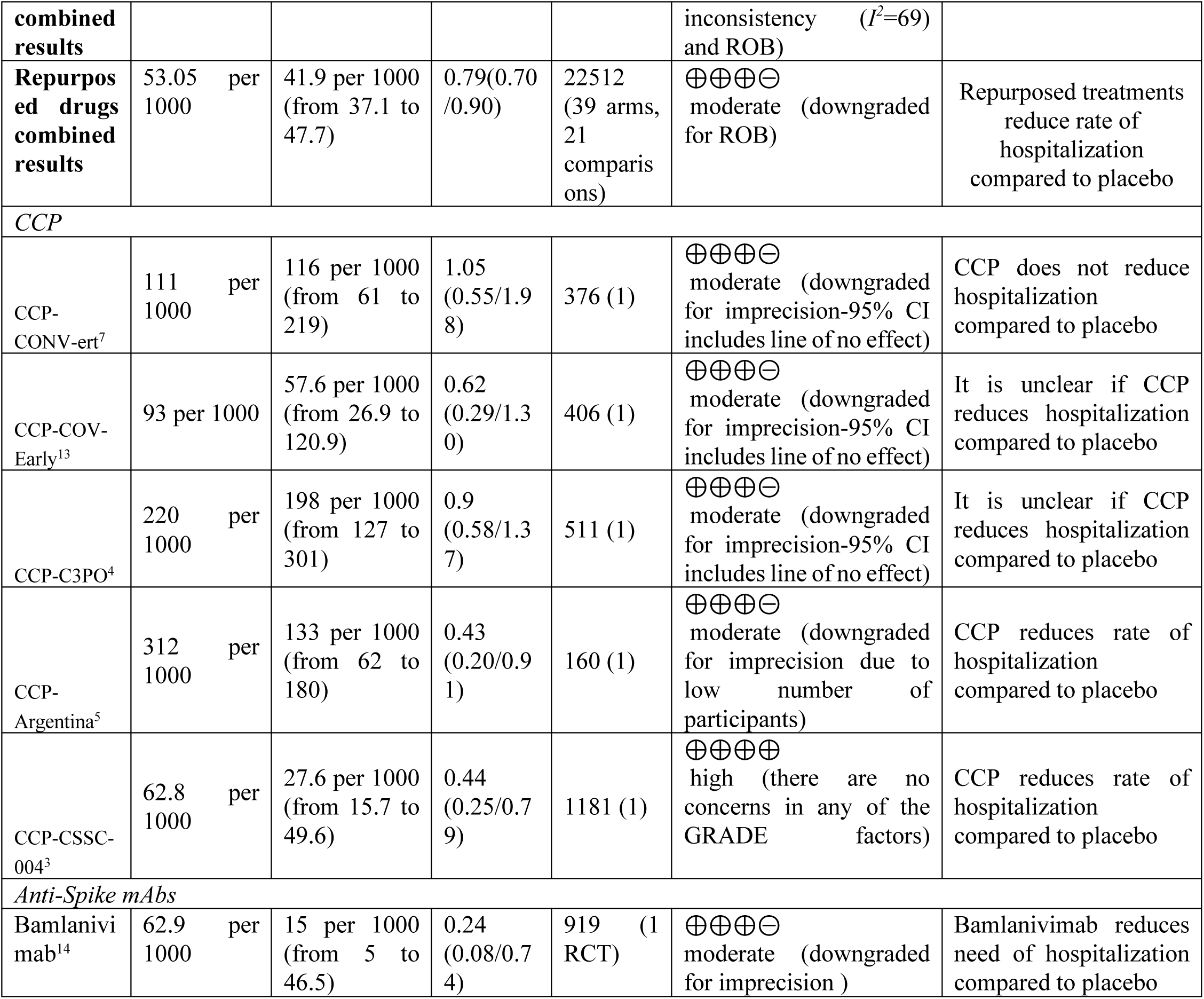
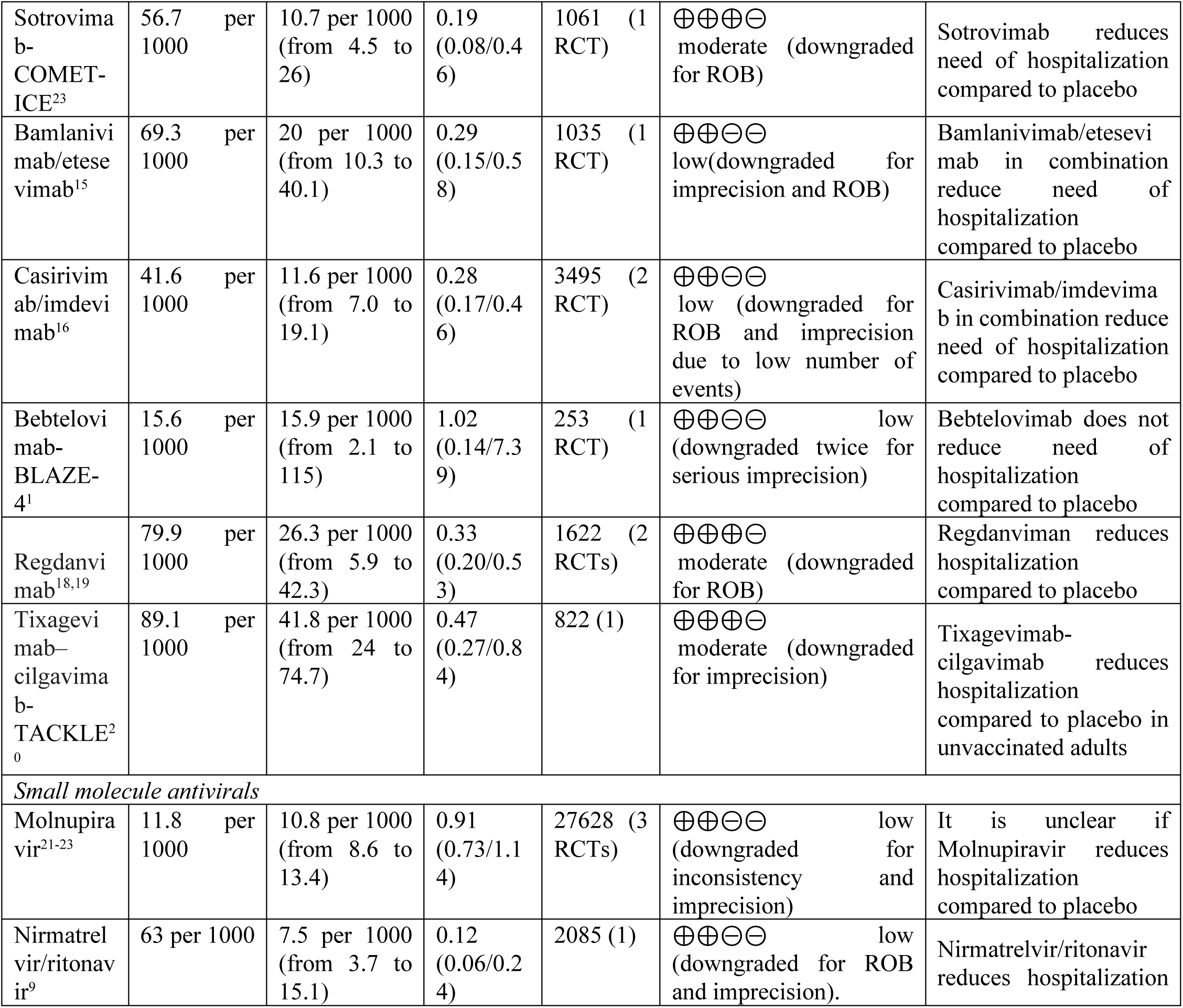
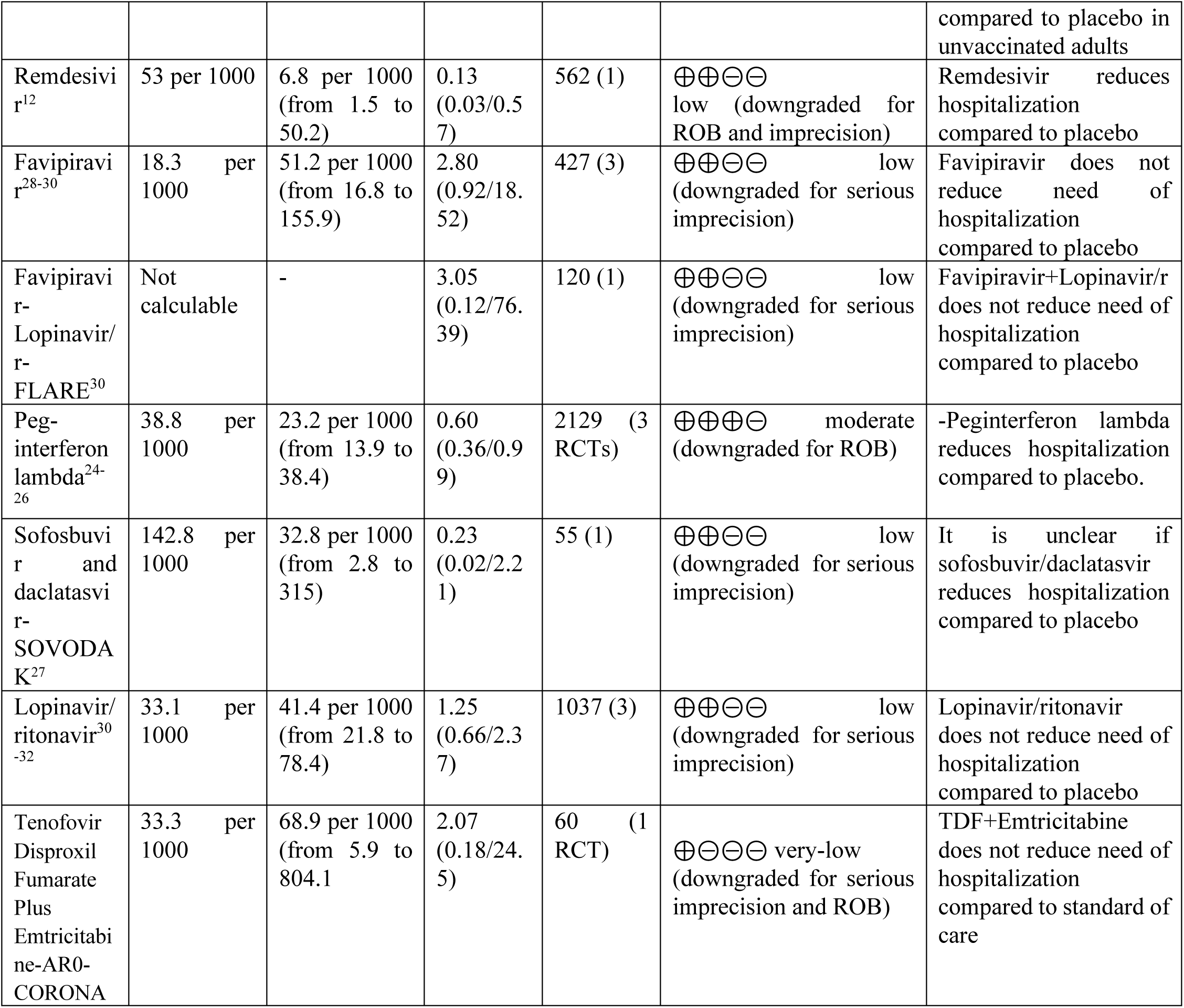
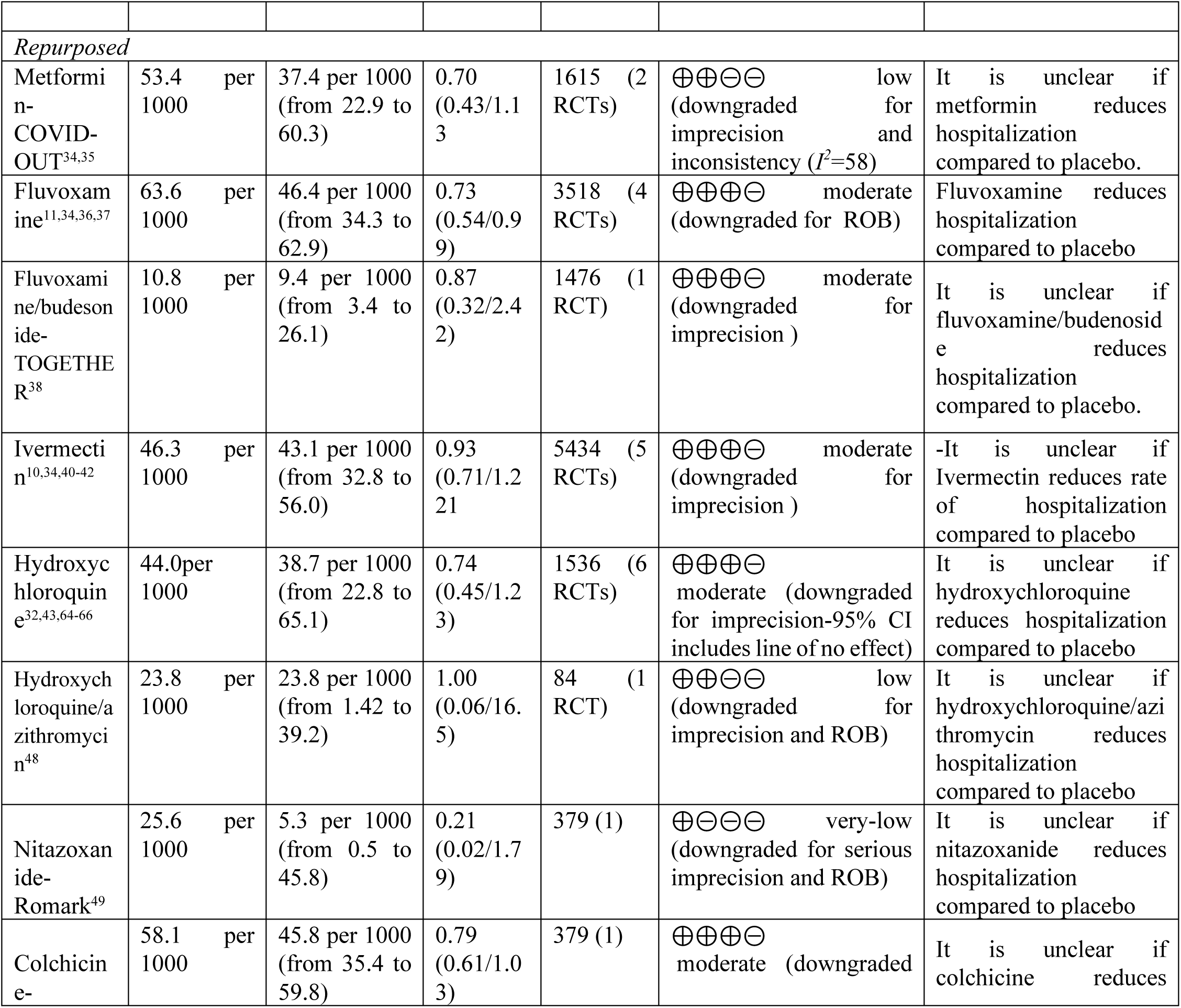
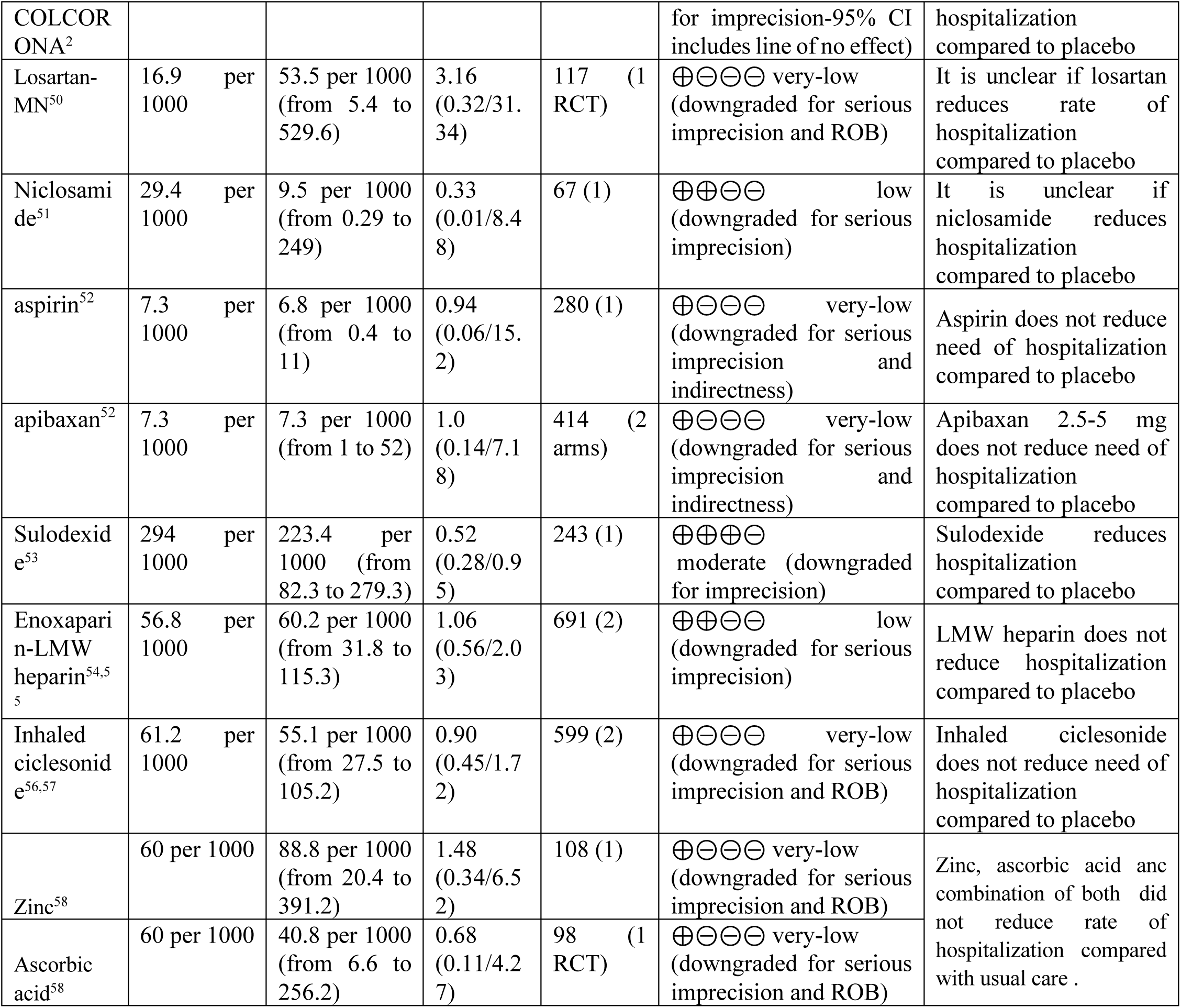
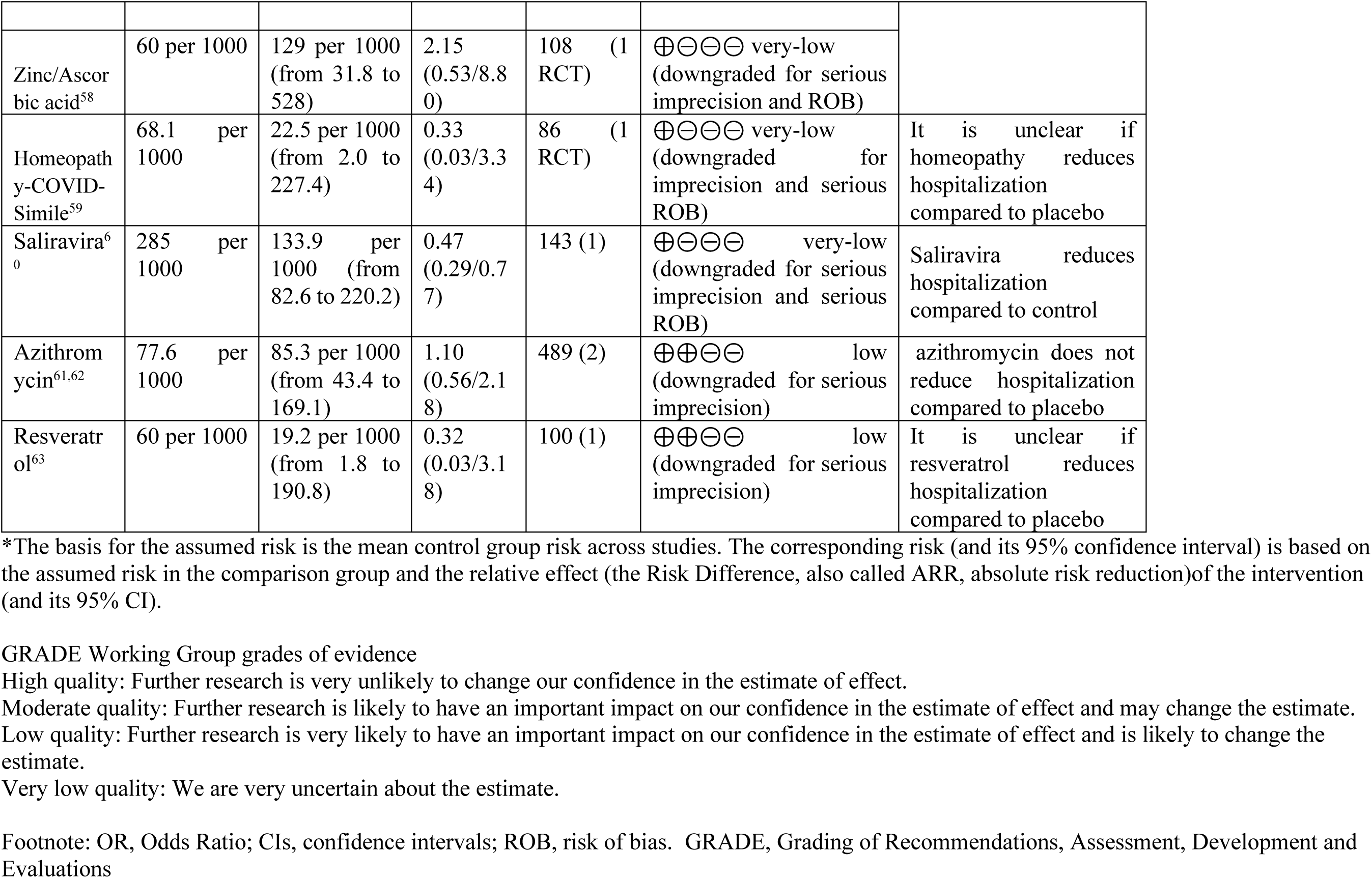
Summary of findings table GRADE evaluation by RCT. Patient or population: COVID-19 outpatients Settings: Ambulatory patients with COVID-19 Intervention: COVID-19 convalescent plasma, anti-Spike mAbs, small molecule antivirals and repurposed drugs Comparison: standard of care, placebo

**Appendix Table 3:** Hospitalized Odds Ratio statistics.

| **Study** | **Hospitaliza**<br>**tion Odds**<br>**ratio** | **Hospitaliza**<br>**tion 95 CI**<br>**low** | **Hospitaliza**<br>**tion 95 CI**<br>**high** | **Hospitaliza**<br>**tion**<br>**significance**<br>**(p)** | **Hospitaliza**<br>**tion**<br>**statistic**<br>**z** |
| --- | --- | --- | --- | --- | --- |
| **Total CCP** | **0.69** | **0.53** | **0.90** | **0.0035** | **2.697265** |
| **Total mAb** | **0.31** | **0.24** | **0.4** | **P<0.001** | **9.29792** |
| **Total antivirals** | **0.78** | **0.45** | **1.33** | **P<0.001** | **3.9493** |
| **Total repurposed** | **0.82** | **0.72** | **0.93** | **P<0.001** | **3.74264** |
| **Total** | **0.67** | **0.57** | **0.80** | **P<0.001** | **9.01288** |
| CCP-CONV-ert<sup>7</sup> | 1.05 | 0.56 | 1.99 | 0.4356 | 0.162033 |
| CCP-COV-Early<sup>13</sup> | 0.61 | 0.29 | 1.30 | 0.1021 | 1.269694 |
| CCP-C3PO<sup>4</sup> | 0.90 | 0.59 | 1.37 | 0.3078 | 0.502004 |
| CCP-Argentina<sup>5</sup> | 0.43 | 0.20 | 0.91 | 0.0140 | 2.197789 |
| CCP-CSSC-004<sup>3</sup> | 0.44 | 0.25 | 0.79 | 0.0031 | 2.737543 |
| CCP-Argentina (high titer)<sup>5</sup> | 0.20 | 0.06 | 0.71 | 0.0132 | 2.478 |
| CCP-CSSC-004 (<= 5 days)<br><sup>3</sup> | 0.18 | 0.07 | 0.49 | 0.0007 | 3.38 |
| Bamlanivimab-BLAZE-1<sup>14</sup> | 0.24 | 0.08 | 0.74 | 0.0066 | 2.479993 |
| Sotrovimab-COMET-ICE<sup>8</sup> | 0.19 | 0.08 | 0.46 | 0.0001 | 3.663844 |
| Bamlanivimab/etesevimab-BLAZE-1<sup>15</sup> | 0.29 | 0.15 | 0.58 | 0.0002 | 3.534471 |
| Casirivimab/imdevimab-REGEN-COV Ph 3<sup>16</sup> | 0.28 | 0.16 | 0.47 | 0.0000 | 4.734662 |
| Casirivimab/imdevimab-REGEN-COV Ph 1/2<sup>17</sup> | 0.30 | 0.07 | 1.25 | 0.0484 | 1.660556 |
| Bebtelovimab-BLAZE-4<sup>1</sup> | 1.02 | 0.14 | 7.39 | 0.4905 | 0.023906 |
| Regdanvimab-CT-P59<sup>18</sup> | 0.49 | 0.19 | 1.27 | 0.0716 | 1.463817 |
| Regdanvimab-CT-P59-2<sup>19</sup> | 0.29 | 0.16 | 0.52 | 0.00001 | 4.22581 |
| Tixagevimab-cilgavimab-TACKLE<sup>20</sup> | 0.47 | 0.26 | 0.84 | 0.0057 | 2.528551 |
| Molnupiravir-MOVE-OUT<sup>21</sup> | 0.67 | 0.46 | 0.99 | 0.0223 | 2.00829 |
| Molnupiravir-PANORAMIC<sup>22</sup> | 1.07 | 0.81 | 1.42 | 0.3156 | 0.479984 |
| Molnupiravir-Aurobindo<sup>23</sup> | NA | NA | NA | NA | NA |
| Nirmatrelvir/ritonavir-EPIC-HR<sup>9</sup> | 0.12 | 0.06 | 0.24 | 0.0000 | 5.731691 |
| Remdesivir-PINETREE<sup>12</sup> | 0.13 | 0.03 | 0.57 | 0.0034 | 2.703067 |
| Interferon Lambda-TOGETHER<sup>24</sup> | 0.47 | 0.29 | 0.77 | 0.0011 | 3.054966 |
| Interferon Lambda- ILIAD<sup>25</sup> | 1.00 | 0.06 | 16.76 | 0.5000 | 0 |
| Interferon Lambda-COVID-Lambda<sup>26</sup> | 1.00 | 0.14 | 7.34 | 0.5000 | 0 |
| Sofosbuvir and daclatasvir-SOVODAK<sup>27</sup> | 0.23 | 0.02 | 2.21 | 0.1018 | 1.271414 |
| Favipavir-Avi-Mild-19<sup>28</sup> | 3.31 | 0.65 | 16.76 | 0.0739 | 1.44706 |
| Favipiravir-Iran<sup>29</sup> | 2.18 | 0.37 | 12.65 | 0.19324 | 0.86602 |
| Favipiravir-FLARE<sup>30</sup> | 3.1 | 0.12 | 77.71 | 0.24541 | 0.68902 |
| Favipiravir/Lopinavir/Ritonavir-FLARE<sup>30</sup> | 3 | 0.12 | 75.11 | 0.25187 | 0.66863 |
| Lopinavir/Ritonavir-FLARE<sup>30</sup> | 3.05 | 0.12 | 76.39 | 0.24865 | 0.67874 |
| Lopinavir/ritonavir-TREAT NOW<sup>31</sup> | 1.21 | 0.4 | 3.64 | 0.37059 | 0.3303 |
| Lopinavir/ritonavir-TOGETHER<sup>32</sup> | 1.20 | 0.53 | 2.69 | 0.3333 | 0.430926 |
| Tenofovir Disproxil Fumarate Plus Emtricitabine-AR0-CORONA<sup>33</sup> | 2.07 | 0.18 | 24.15 | 0.28057 | 0.58116 |
| Metformin-COVID-OUT<sup>34</sup> | 0.42 | 0.18 | 0.96 | 0.0198 | 2.057 |
| Metformin -TOGETHER<sup>35</sup> | 0.94 | 0.51 | 1.71 | 0.41626 | 0.21147 |
| Fluvoxamine-TOGETHER<sup>11</sup> | 0.77 | 0.56 | 1.05 | 0.0505 | 1.640023 |
| Fluvoxamine-STOP COVID<sup>36</sup> | 0.06 | 0.00 | 1.15 | 0.0310 | 1.866043 |
| Fluvoxamine-COVID-OUT<sup>34</sup> | 1.18 | 0.36 | 3.91 | 0.3935 | 0.270145 |
| Fluvoxamine ACTIV-6<sup>37</sup> | 0.45 | 0.04 | 5 | 0.25869 | 0.64738 |
| Fluvoxamine/budesonide-TOGETHER<sup>38</sup> | 0.87 | 0.32 | 2.42 | 0.39769 | 0.25933 |
| Ivermectin-TOGETHER<sup>39</sup> | 0.81 | 0.59 | 1.11 | 0.09724 | 1.29742 |
| Ivermectin-COVID-OUT<sup>34</sup> | 0.76 | 0.20 | 2.85 | 0.3414 | 0.408715 |
| Ivermectin Iran<sup>40</sup> | 1.46 | 0.71 | 2.96 | 0.1507 | 1.033341 |
| Ivermectin-ACTIV-6<sup>41</sup> | 1.05 | 0.43 | 2.61 | 0.4553 | 0.112309 |
| Ivermectin high dose-ACTIV-6<sup>42</sup> | 2.52 | 0.49 | 13.04 | 0.13512 | 1.10252 |
| Hydroxychloroquine-TOGETHER<sup>32</sup> | 0.76 | 0.30 | 1.93 | 0.2840 | 0.570928 |
| Hydroxychloroquine-COVID-19 PEP<sup>43</sup> | 0.49 | 0.16 | 1.45 | 0.0971 | 1.298164 |
| Hydroxychloroquine -AH COVID-19<sup>44</sup> | 3.14 | 0.17 | 59.70 | 0.2232 | 0.761332 |
| Hydroxychloroquine-BCN PEP-CoV-2<sup>45</sup> | 0.83 | 0.32 | 2.13 | 0.3486 | 0.389184 |
| Hydroxychloroquine-BMG<sup>46</sup> | 0.69 | 0.18 | 2.65 | 0.2945 | 0.54027 |
| Hydroxychloroquine-Utah<sup>47</sup> | 1.77 | 0.51 | 6.19 | 0.18429 | 0.89915 |
| Hydroxychloroquine/Azithromycin-Brazil<sup>48</sup> | 1 | 0.06 | 16.5 | 0.5 | 0 |
| Nitazoxanide-Romark<sup>49</sup> | 0.21 | 0.02 | 1.79 | 0.0766 | 1.428571 |
| Colchicine-COLCORONA<sup>2</sup> | 0.79 | 0.61 | 1.03 | 0.0407 | 1.742726 |
| Losartan-MN<sup>50</sup> | 3.16 | 0.32 | 31.34 | 0.16245 | 0.98443 |
| Niclosamide<sup>51</sup> | 0.33 | 0.01 | 8.48 | 0.2529 | 0.665353 |
| Aspirin-ACTIV-4B<sup>52</sup> | 0.94 | 0.06 | 15.24 | 0.4838 | 0.040562 |
| 2.5-mg apixaban-ACTIV-4B<sup>52</sup> | 1.01 | 0.06 | 16.27 | 0.4979 | 0.005238 |
| 5-mg apixaban ACTIV-4B<sup>52</sup> | 1.91 | 0.17 | 21.36 | 0.2988 | 0.527902 |
| Sulodexide<sup>53</sup> | 0.52 | 0.28 | 0.95 | 0.0167 | 2.128119 |
| Enoxaparin-ETHIC<sup>54</sup> | 1.10 | 0.47 | 2.56 | 0.4155 | 0.213485 |
| Enoxaparin-OVID<sup>55</sup> | 1.02 | 0.38 | 2.76 | 0.4862 | 0.034489 |
| Inhaled Ciclesonide-COVIS<sup>56</sup> | 0.48 | 0.12 | 1.87 | 0.14459 | 1.05994 |
| Inhaled ciclesonide-COVERAGE<sup>57</sup> | 1.15 | 0.51 | 2.63 | 0.3659 | 0.34277 |
| Zinc<sup>58</sup> | 1.48 | 0.36 | 6.52 | 0.30296 | 0.5159 |
| Ascorbic acid<sup>58</sup> | 0.68 | 0.11 | 4.27 | 0.34085 | 0.41015 |
| Zinc/Ascorbic acid<sup>58</sup> | 2.15 | 0.53 | 8.8 | 0.1435 | 1.06472 |
| Homeopathy-COVID-Simile<sup>59</sup> | 0.33 | 0.03 | 3.34 | 0.17503 | 0.93449 |
| Saliravira<sup>60</sup> | 0.01 | 0.00 | 0.24 | 0.0016 | 2.9467 |
| Azithromycin-Atomic2<sup>61</sup> | 0.88 | 0.42 | 1.84 | 0.3694 | 0.333472 |
| Azithromycin-ACTION<sup>62</sup> | 6.62 | 0.36 | 121.45 | 0.1015 | 1.272994 |
| Resveratrol<sup>63</sup> | 0.32 | 0.03 | 3.18 | 0.1654 | 0.972432 |

**Appendix Table 4:** Deaths during RCTs. Cumulatively, the CCP RCTs noted 10 deaths in the control arm versus 8 in CCP arm. The anti-Spike mAbs RCTs had 21 total deaths among controls and 4 in the intervention arm. The total for all small molecule antiviral RCTs was 28 in the controls and 7 in the interventions. The repurposed drugs RCTs recorded 72 deaths in the control groups and 53 in the intervention groups.

| Study | ARR % | RRR % | 95% CI ARR | 95% CI RRR | Odds ratio | 95 CI low | 95 CI high | z statistic | significance (p) |
| --- | --- | --- | --- | --- | --- | --- | --- | --- | --- |
| Total CCP | 0.15 | 20.2 | (-0.48, 0.78) | (-101.5, 68.4) | 0.8 | 0.31 | 2.02 | 0.47848 | 0.31616 |
| Total mAb | 0.45 | 80.8 | (0.21, 0.7) | (49.4, 92.7) | 0.19 | 0.07 | 0.5 | 3.3459 | 0.00041 |
| Total antivirals | 0.16 | 87.1 | (0.1, 0.23) | (63.4, 95.4) | 0.13 | 0.05 | 0.37 | 3.85181 | 0.00006 |
| Total repurposed | 0.16 | 21.8 | (-0.1, 0.4) | (-8.3, 43.5) | 0.78 | 0.56 | 1.08 | 1.4795 | 0.0695 |
| Total | 0.19 | 44.7 | (0.11, 0.28) | (27.6, 57.8) | 0.55 | 0.42 | 0.72 | 4.29906 | 0.00001 |
| Study | Deaths control | Total control | Deaths intervention | Total intervention | Total both arms | % death control | % death intervention |  |  |
| **Total CCP** | 10 | 1315 | 8 | 1319 | 2634 | 0.76 | 0.61 |  |  |
| **Total mAb** | 23 | 4102 | 5 | 4634 | 8736 | 0.56 | 0.11 |  |  |
| **Total antivirals** | 31 | 17073 | 4 | 17031 | 34104 | 0.18 | 0.02 |  |  |
| **Total repurposed** | 81 | 11109 | 65 | 11396 | 22505 | 0.73 | 0.57 |  |  |
| **Total** | 131 | 29680 | 72 | 30363 | 60043 | 0.44 | 0.24 |  |  |
| CCP-CONV-ert<sup>7</sup> | 2 | 188 | 0 | 188 | 376 | 1.06 | 0 |  |  |
| CCP-COV-Early<sup>13</sup> | 0 | 204 | 1 | 202 | 406 | 0 | 0.5 |  |  |
| CCP-C3PO<sup>4</sup> | 1 | 254 | 5 | 257 | 511 | 0.39 | 1.95 |  |  |
| CCP-Argentina<sup>5</sup> | 4 | 80 | 2 | 80 | 160 | 5 | 2.5 |  |  |
| CCP-CSSC-004<sup>3</sup> | 3 | 589 | 0 | 592 | 1181 | 0.51 | 0 |  |  |
| Bamlanivimab-BLAZE-1<sup>14</sup> | 0 | 143 | 0 | 309 | 452 | 0 | 0 |  |  |
| Sotrovimab-COMET-ICE<sup>8</sup> | 2 | 529 | 0 | 528 | 1057 | 0.38 | 0 |  |  |
| Bamlanivimab/etesevimab-BLAZE-1<sup>15</sup> | 10 | 517 | 0 | 518 | 1035 | 1.93 | 0 |  |  |
| Casirivimab/<br>imdevimab-REGEN-COV Ph 3<sup>16</sup> | 3 | 1341 | 1 | 1355 | 269<br>6 | 0.22 | 0.07 |  |  |
| Casirivimab/<br>imdevimab-REGEN-COV Ph 1/2<sup>17</sup> | 0 | 266 | 0 | 533 | 799 | 0 | 0 |  |  |
| Bebtelovimab-BLAZE-<br>4<sup>1</sup> | 0 | 128 | 0 | 125 | 253 | 0 | 0 |  |  |
| Regdanvimab-CT-P59<sup>18</sup> | 0 | 104 | 0 | 203 | 307 | 0 | 0 |  |  |
| Regdanvimab-CT-P59-<br>2<sup>19</sup> | 2 | 659 | 1 | 656 | 131<br>5 | 0.3 | 0.15 |  |  |
| Tixagevimab–<br>cilgavimab-TACKLE<sup>20</sup> | 6 | 415 | 3 | 407 | 822 | 1.45 | 0.74 |  |  |
| Molnupiravir-MOVE-OUT<sup>21</sup> | 9 | 699 | 1 | 709 | 140<br>8 | 1.29 | 0.14 |  |  |
| Molnupiravir-PANORAMIC<sup>22</sup> | 5 | 1248<br>4 | 2 | 12516 | 250<br>00 | 0.04 | 0.02 |  |  |
| Molnupiravir-Aurobindo<sup>23</sup> | 0 | 610 | 0 | 610 | 122<br>0 | 0 | 0 |  |  |
| Nirmatrelvir/ritonavir-EPIC-HR<sup>9</sup> | 12 | 1046 | 0 | 1039 | 208<br>5 | 1.15 | 0 |  |  |
| Remdesivir-<br>PINETREE<sup>12</sup> | 1 | 283 | 0 | 279 | 562 | 0.35 | 0 |  |  |
| Interferon Lambda-TOGETHER<sup>24</sup> | 4 | 1018 | 1 | 931 | 194<br>9 | 0.4 | 0.11 |  |  |
| Interferon Lambda-ILIAD<sup>25</sup> | 0 | 30 | 0 | 30 | 60 | 0 | 0 |  |  |
| Interferon Lambda-COVID-Lambda<sup>26</sup> | 0 | 60 | 0 | 60 | 120 | 0 | 0 |  |  |
| Sofosbuvir and<br>daclatasvir-SOVODAK<sup>27</sup> | 0 | 28 | 0 | 27 | 55 | 0 | 0 |  |  |
| Favipavir-Avi-Mild-19<sup>28</sup> | 0 | 119 | 0 | 112 | 231 | 0 | 0 |  |  |
| Favipiravir-Iran<sup>29</sup> | 0 | 39 | 0 | 38 | 77 | 0 | 0 |  |  |
| Favipiravir-FLARE<sup>30</sup> | 0 | 60 | 0 | 59 | 119 | 0 | 0 |  |  |
| Favipiravir/Lopinavir/Ri-<br>tonavir-FLARE<sup>30</sup> | 0 | 60 | 0 | 61 | 121 | 0 | 0 |  |  |
| Lopinavir/Ritonavir-FLARE<sup>30</sup> | 0 | 60 | 0 | 60 | 120 | 0 | 0 |  |  |
| Lopinavir/ritonavir-TREAT NOW<sup>31</sup> | 0 | 220 | 0 | 226 | 446 | 0 | 0 |  |  |
| Lopinavir/ritonavir-TOGETHER<sup>32</sup> | 0 | 227 | 0 | 244 | 471 | 0 | 0 |  |  |
| Tenofovir Disproxil<br>Fumarate Plus<br>Emtricitabine-AR0-CORONA<sup>33</sup> | 0 | 30 | 0 | 30 | 60 | 0 | 0 |  |  |
| Metformin-COVID-OUT<sup>34</sup> | 1 | 601 | 1 | 596 | 119<br>7 | 0.17 | 0.17 |  |  |
| Metformin - TOGETHER<sup>35</sup> | 9 | 203 | 7 | 215 | 418 | 4.4 | 3.2 |  |  |
| Fluvoxamine-TOGETHER<sup>11</sup> | 25 | 756 | 17 | 741 | 149<br>7 | 3.31 | 2.29 |  |  |
| Fluvoxamine-STOP COVID<sup>36</sup> | 0 | 72 | 0 | 80 | 152 | 0 | 0 |  |  |
| Fluvoxamine-COVID-OUT<sup>34</sup> | 0 | 293 | 0 | 299 | 592 | 0 | 0 |  |  |
| Fluvoxamine ACTIV-6<sup>37</sup> | 0 | 599 | 0 | 662 | 126<br>1 | 0 | 0 |  |  |
| Fluvoxamine/budesonide-TOGETHER<sup>38</sup> | 0 | 738 | 1 | 738 | 147<br>6 | 0 | 0.14 |  |  |
| Ivermectin-TOGETHER<sup>39</sup> | 24 | 675 | 21 | 674 | 134<br>9 | 3.56 | 3.12 |  |  |
| Ivermectin-COVID-OUT<sup>34</sup> | 0 | 356 | 1 | 374 | 730 | 0 | 0.27 |  |  |
| Ivermectin Iran<sup>40</sup> | 1 | 281 | 1 | 268 | 549 | 0.36 | 0.37 |  |  |
| Ivermectin-ACTIV-6<sup>41</sup> | 0 | 774 | 1 | 817 | 159<br>1 | 0 | 0.12 |  |  |
| Ivermectin high dose-ACTIV-6<sup>42</sup> | 0 | 604 | 1 | 602 | 120<br>6 | 0 | 0.17 |  |  |
| Hydroxychloroquine-TOGETHER<sup>32</sup> | 1 | 227 | 0 | 214 | 441 | 0.44 | 0 |  |  |
| Hydroxychloroquine-COVID-19 PEP<sup>43</sup> | 1 | 211 | 1 | 212 | 423 | 0.47 | 0.47 |  |  |
| Hydroxychloroquine - AH COVID-19<sup>44</sup> | 0 | 37 | 0 | 111 | 148 | 0 | 0 |  |  |
| Hydroxychloroquine-BCN PEP-CoV-2<sup>45</sup> | 0 | 157 | 0 | 136 | 293 | 0 | 0 |  |  |
| Hydroxychloroquine-BMG<sup>46</sup> | 0 | 83 | 0 | 148 | 231 | 0 | 0 |  |  |
| Hydroxychloroquine-Utah<sup>47</sup> | 0 | 151 | 0 | 152 | 303 | 0 | 0 |  |  |
| Hydroxychloroquine/Azithromycin-Brazil<sup>48</sup> | 0 | 42 | 0 | 42 | 84 | 0 | 0 |  |  |
| Nitazoxanide-Romark<sup>49</sup> | 0 | 195 | 0 | 184 | 379 | 0 | 0 |  |  |
| Colchicine-COLCORONA<sup>2</sup> | 9 | 2253 | 5 | 2235 | 448<br>8 | 0.4 | 0.22 |  |  |
| Losartan-MN<sup>50</sup> | 0 | 59 | 0 | 58 | 117 | 0 | 0 |  |  |
| Niclosamide<sup>51</sup> | 0 | 34 | 0 | 33 | 67 | 0 | 0 |  |  |
| Aspirin-ACTIV-4B<sup>52</sup> | 0 | 136 | 0 | 144 | 280 | 0 | 0 |  |  |
| 2.5-mg apixaban-ACTIV-4B<sup>52</sup> | 0 | 136 | 0 | 135 | 271 | 0 | 0 |  |  |
| 5-mg apixaban ACTIV-4B<sup>52</sup> | 0 | 136 | 0 | 143 | 279 | 0 | 0 |  |  |
| Sulodexide<sup>53</sup> | 7 | 119 | 3 | 124 | 243 | 5.88 | 2.42 |  |  |
| Enoxaparin-ETHIC<sup>54</sup> | 0 | 114 | 1 | 105 | 219 | 0 | 0.95 |  |  |
| Enoxaparin-OVID<sup>55</sup> | 0 | 238 | 0 | 234 | 472 | 0 | 0 |  |  |
| Inhaled Ciclesonide-COVIS<sup>56</sup> | 0 | 203 | 0 | 197 | 400 | 0 | 0 |  |  |
| Inhaled ciclesonide-COVERAGE<sup>57</sup> | 2 | 107 | 0 | 110 | 217 | 1.87 | 0 |  |  |
| Zinc<sup>58</sup> | 0 | 50 | 0 | 58 | 108 | 0 | 0 |  |  |
| Ascorbic acid<sup>58</sup> | 0 | 50 | 1 | 48 | 98 | 0 | 2.08 |  |  |
| Zinc/Ascorbic acid<sup>58</sup> | 0 | 50 | 2 | 58 | 108 | 0 | 3.45 |  |  |
| Homeopathy-COVID-Simile<sup>59</sup> | 0 | 44 | 0 | 42 | 86 | 0 | 0 |  |  |
| Saliravira<sup>60</sup> | 0 | 56 | 0 | 87 | 143 | 0 | 0 |  |  |
| Azithromycin-Atomic2<sup>61</sup> | 1 | 147 | 1 | 145 | 292 | 0.68 | 0.69 |  |  |
| Azithromycin-ACTION<sup>62</sup> | 0 | 72 | 0 | 125 | 197 | 0 | 0 |  |  |
| Resveratrol<sup>63</sup> | 0 | 50 | 0 | 50 | 100 | 0 | 0 |  |  |

